# Comparative survey-based study of non-invasive saliva collection devices

**DOI:** 10.1101/2023.10.31.23297784

**Authors:** Yeokyoung (Anne) Kil, Ali S. Booeshaghi, Lior S. Pachter

## Abstract

**Background:** We compared five saliva collection devices on their saliva collection efficiency, instruction reading rate, user difficulty ratings, and leakage of saliva, all of which are important factors in safe, easy, and efficient saliva collection. The devices evaluated were: Salivette (swab), SuperSAL (swab), SalivaBio Passive Drool, Medschenker Saliva Collection Kit (funnel), and cryovial with funnel used in SwabSeq COVID-19 surveillance tests.

**Methods:** 56 individuals used five devices in randomized orders by first reading the device’s instruction manual while timed, then self-collecting saliva while timed, to measure the instruction reading rate and saliva collection rate, respectively. For each device, users were asked about the difficulties of instructions; assembly; and saliva collection, and whether there was leakage of saliva. Lastly, unstimulated and stimulated saliva production (=flow) rates for each user were measured. The saliva collection and instruction reading rates were normalized by the individual’s base saliva flow rate and base reading rate. The rates and difficulty ratings for devices were compared using permutation tests and one-way ANOVA.

**Results:** Salivette had the highest average saliva collection rate and SuperSAL had the lowest. For the instruction reading rate, Medschenker’s funnel device had the highest average and Salivette had the lowest. While all devices showed saliva leakage, passive drool had the highest fraction of leakages and the Medschenker device the lowest. Users found the instructions for Salivette the hardest and those for SwabSeq the easiest. Users found the assembly for Medschenker to be easiest and that for SuperSAL to be hardest. Users rated Salivette easiest to collect saliva with, and SuperSAL most difficult.

**Conclusions:** Medschenker performed well on most qualitative and quantitative metrics while SuperSAL did not perform as well. However, no single saliva collection method or device satisfies all requirements of an ideal device. A device that allows for efficient saliva collection, easy usage, and safe saliva collection without leakage could greatly help standardize saliva collection.

## Background

Saliva is an established biofluid for medical diagnostics. For example, saliva is used to diagnose Cushing Syndrome, an endocrine disorder in which the body produces too much cortisol (1). Saliva has also been used for point-of-care testing; In 2012, the FDA approved the OraQuick In-Home HIV test, which detects antibodies to HIV Types 1 and 2 in saliva (2). In the course of the COVID-19 pandemic, many saliva-based diagnostic tests for SARS-CoV-2 were developed and utilized to test individuals for medical and public health purposes. These include nucleic acid amplification tests (NAAT) like PCR tests, and some antigen tests.

Saliva has many characteristics that make it an attractive sample type for diagnosis. The biggest advantage in using saliva for diagnosis is that although some gland collection techniques are invasive, it is straightforward to collect whole saliva noninvasively (3). Saliva is easier to collect than blood, and is also more readily produced than sweat or urine, which are other biofluids that can be collected noninvasively. Saliva samples are stable in room temperature and in higher temperatures without preservation buffers (4), a feature which facilitates flexibility in transportation of samples.

There are many different methods and devices for collecting saliva non-invasively. The methods can be broadly classified based on three distinct collection mechanisms: swabs, funnels, and passive drool. Swabs refer to absorbent materials like cotton or synthetic rolls that are placed inside the mouth to passively absorb saliva. Funnels can either be detachable or integrated with the collection tube, and help guide the expectorated saliva into the tube. Passive drooling refers to a method in which the user lets saliva naturally pool in their mouths without actively gathering saliva with their tongues, with collection proceeding via the pooled saliva gravitationally flowing into a collection tube. Each mechanism has its own strengths and weaknesses, and one mechanism may be more suitable than other mechanisms for certain usage cases of the collected saliva, or for specific demographics. For instance, absorbent swabs may be used for patients who have difficulty spitting, such as pediatric patients (5). However, swabs can interfere with salivary contents, such as hormones (6). The advantages and disadvantages of each saliva collection mechanism are summarized in **Table 1**. The table also contains information about the devices that were used in this study.

**Table 1.**
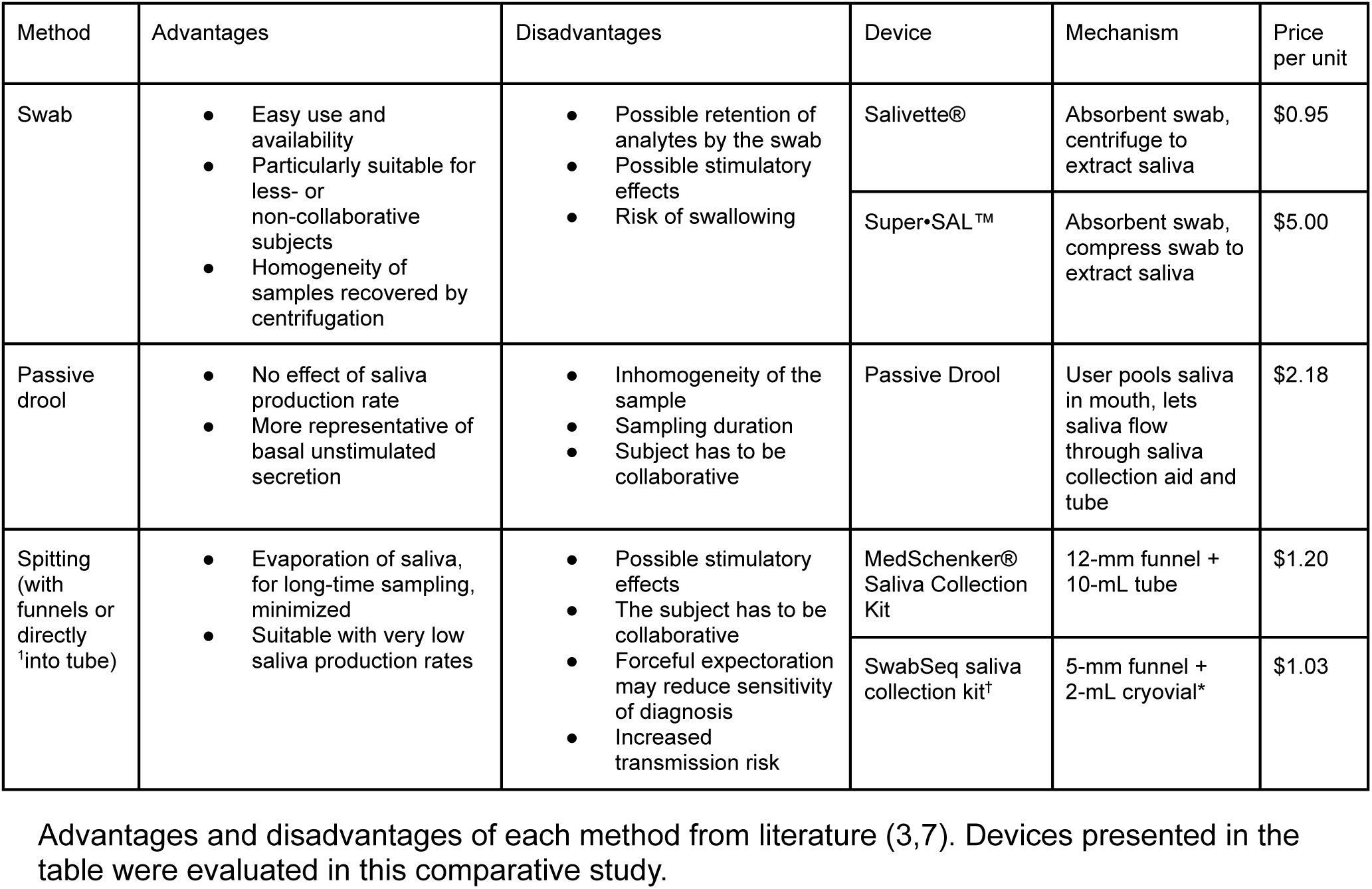
Summary of sample methods and representative devices.

| Method | Advantages | Disadvantages | Device | Mechanism | Price per unit |
| --- | --- | --- | --- | --- | --- |
| Swab | • Easy use and availability<br>• Particularly suitable for less- or non-collaborative subjects<br>• Homogeneity of samples recovered by centrifugation | • Possible retention of analytes by the swab<br>• Possible stimulatory effects<br>• Risk of swallowing | Salivette® | Absorbent swab, centrifuge to extract saliva | \$0.95 |
| | | | Super•SAL™ | Absorbent swab, compress swab to extract saliva | \$5.00 |
| Passive drool | • No effect of saliva production rate<br>• More representative of basal unstimulated secretion | • Inhomogeneity of the sample<br>• Sampling duration<br>• Subject has to be collaborative | Passive Drool | User pools saliva in mouth, lets saliva flow through saliva collection aid and tube | \$2.18 |
| Spitting (with funnels or directly<sup>†</sup> into tube) | • Evaporation of saliva, for long-time sampling, minimized<br>• Suitable with very low saliva production rates | • Possible stimulatory effects<br>• The subject has to be collaborative<br>• Forceful expectoration may reduce sensitivity of diagnosis<br>• Increased transmission risk | MedSchenker® Saliva Collection Kit | 12-mm funnel + 10-mL tube | \$1.20 |
| | | | SwabSeq saliva collection kit<sup>†</sup> | 5-mm funnel + 2-mL cryovial* | \$1.03 |

Advantages and disadvantages of each method from literature (3,7). Devices presented in the table were evaluated in this comparative study.

In addition to the characteristics of the saliva collection mechanism, attributes of each saliva collection device should be considered when choosing a method to sample saliva. Such attributes may include price, availability, downstream processes, and in the case of swabs, the material of the swab. The selection of a device should also be undertaken in the context of its intended application. For instance, in a mass-testing scenario where multiple people are collecting saliva for testing purposes, it may be important to minimize saliva leakage to lower the risks of secondary infections, while ensuring that saliva is collected efficiently to increase the throughput of the tests.

In this survey-based comparative study, we identified five saliva collection devices–two funnel devices, two swab devices, and one passive-drool device (Figure 1). The description of each device is summarized in Table 1. We measured each device’s performance, usability, and hygiene by measuring quantitative and qualitative metrics of during device use, such as the saliva collection rate of the device and the user’s reading rate for the device’s instructions, as well as difficulties of instructions, assembly, and saliva collection. We measured unstimulated saliva flow rates for all subjects, in order to normalize the device performances by individual saliva production (flow) rates. Additionally, to understand the feasibility of using stimulated saliva for saliva diagnostics in the future, we measured each subject’s saliva flow rates under stimulation with water and citric acid solution.

**Figure 1.**
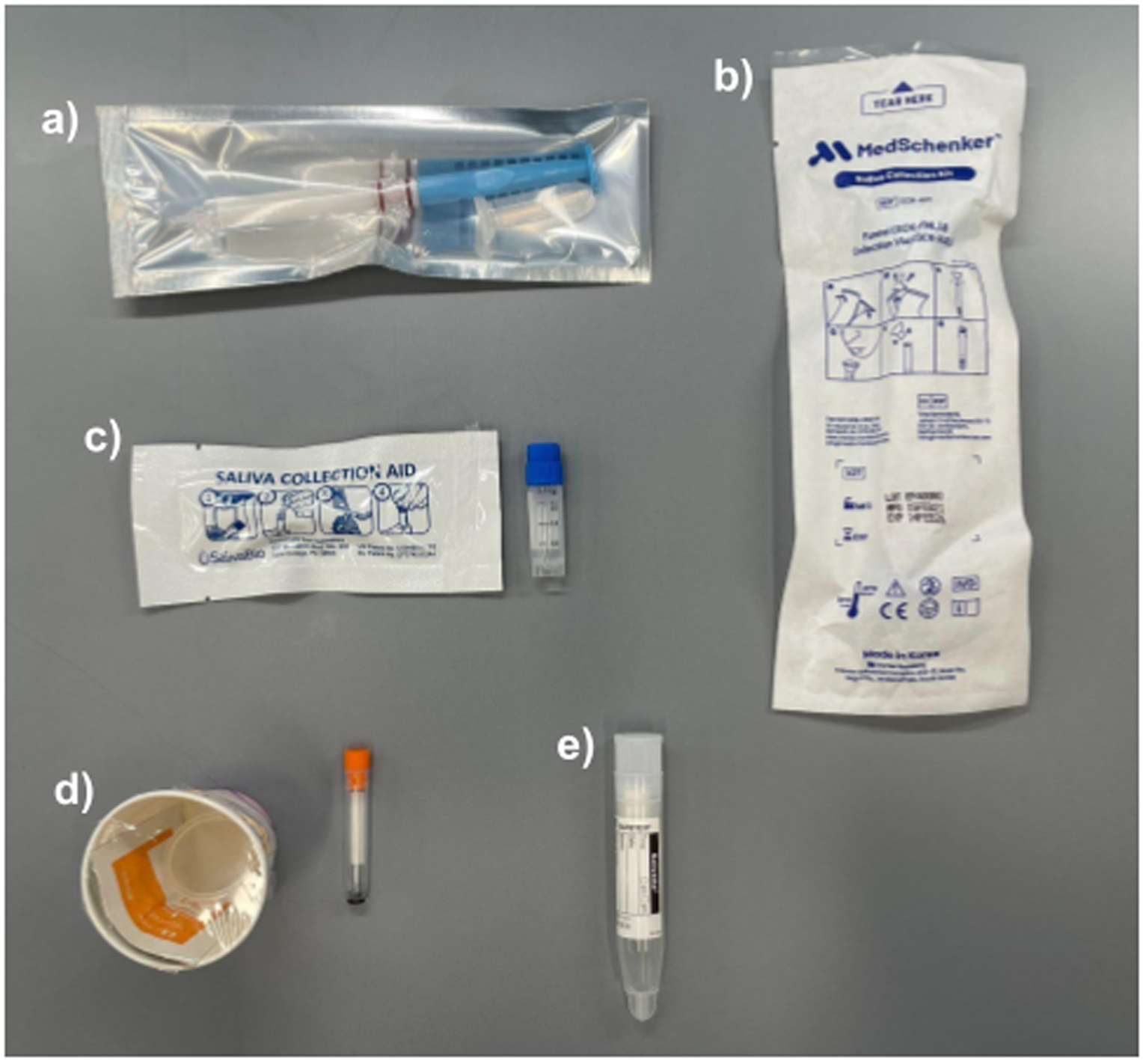
The five saliva collection devices selected for the study. a) SuperSAL, b) MedSchenker, c) Passive Drool, d) SwabSeq, and e) Salivette. More information on each device can be found in **Table 1**.

## Results

Results were first collected for exploratory samples (subjects 1-30) and later for confirmatory samples (31-60). We report the separate results in **<u>Additional File 1</u>** and focus on the combined analysis here. As subjects 57-60 were unable to use the Medschenker device in their sessions due to unforeseen supply issues, they were excluded from the comparison analysis.

### Device comparison

#### Normalized Saliva Collection Rate

The saliva collection rate was measured for each device by timing how long subjects took to collect volumes of saliva specified by the device’s instructions. This number was then normalized by each subject’s unstimulated, base saliva flow rate in order to remove the differences in each person’s saliva production. The normalized rate indicates whether the device facilitated efficient saliva collection or hindered it: a number above 0 indicates that the device had a stimulatory effect on saliva production and thus resulted in efficient saliva collection, and a number around 0 would mean that the device was as efficient in saliva collection as the subject’s natural saliva production. In contrast, a number below 0 indicates that the device either hindered saliva production or resulted in saliva collection that was less efficient than the subject’s natural saliva production. The results are shown in Figure 2 and Table 2.

**Figure 2.**
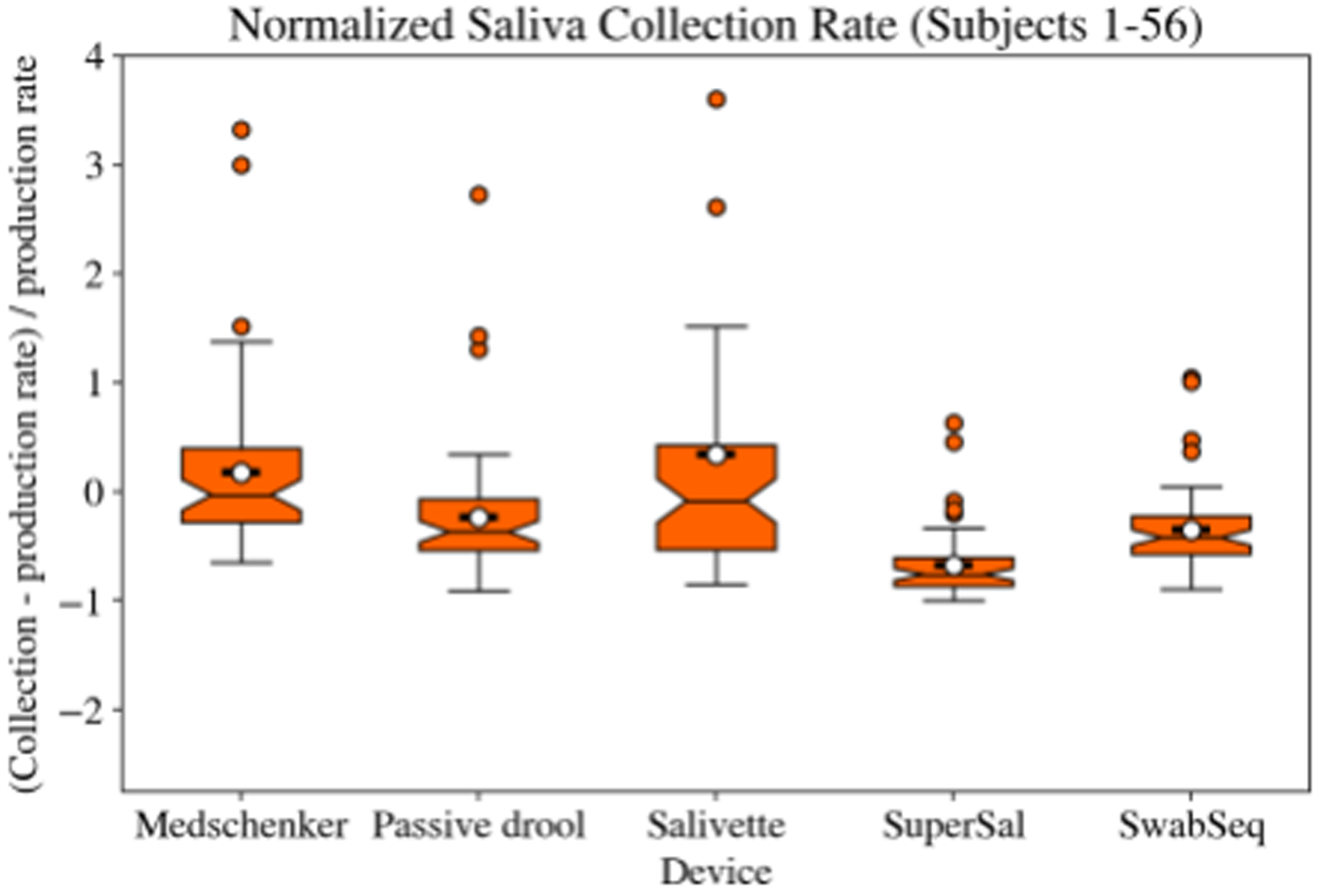
Saliva collection rate of each device, normalized by individual saliva flow rate.

**Table 2.** Average Normalized Saliva Collection Rate (Standard deviation)

| Device | Avg. Saliva Collection Rate (St. dev) [unitless] |
| --- | --- |
| Medschenker | 0.178 (0.747) |
| Passive drool | -0.235 (0.595) |
| Salivette | 0.338 (1.944) |
| SuperSAL | -0.674 (0.311) |
| SwabSeq | -0.351 (0.373) |

The average normalized saliva collection rate was ranked highest to lowest in the order of Salivette, Medschenker, Passive drool, SwabSeq, and SuperSAL. Salivette’s and Medschenker’s average rates were both above 0, while the three other devices had normalized rates below 0. Out of the five devices, Salivette had the highest average and SuperSAL the lowest. Although Salivette had the highest average, it also had the highest standard deviation and highest interquartile range, meaning that the normalized saliva flow rates for the device were more spread out than other devices. SuperSAL had the lowest average but also had the lowest standard deviation and interquartile range.

#### Normalized Instruction Reading Rates

The reading rate for each device’s instruction manual was measured by timing how long subjects took to read the manual and dividing that time by the word count of the manual, prior to the subjects’ using the device. This number was then normalized by each subject’s base reading rate, which was measured while subjects read the consent form document at the beginning of the study. The normalized rate was used as a measure of how easy to read the manuals were. A normalized rate around 0 would indicate that the difficulty of the document is around the same as a plain text document with no jargon. The results are presented in **Figure 3** and **Table 3**. The manuals that were presented to subjects can be found in the repository with the data for the study.

**Figure 3.**
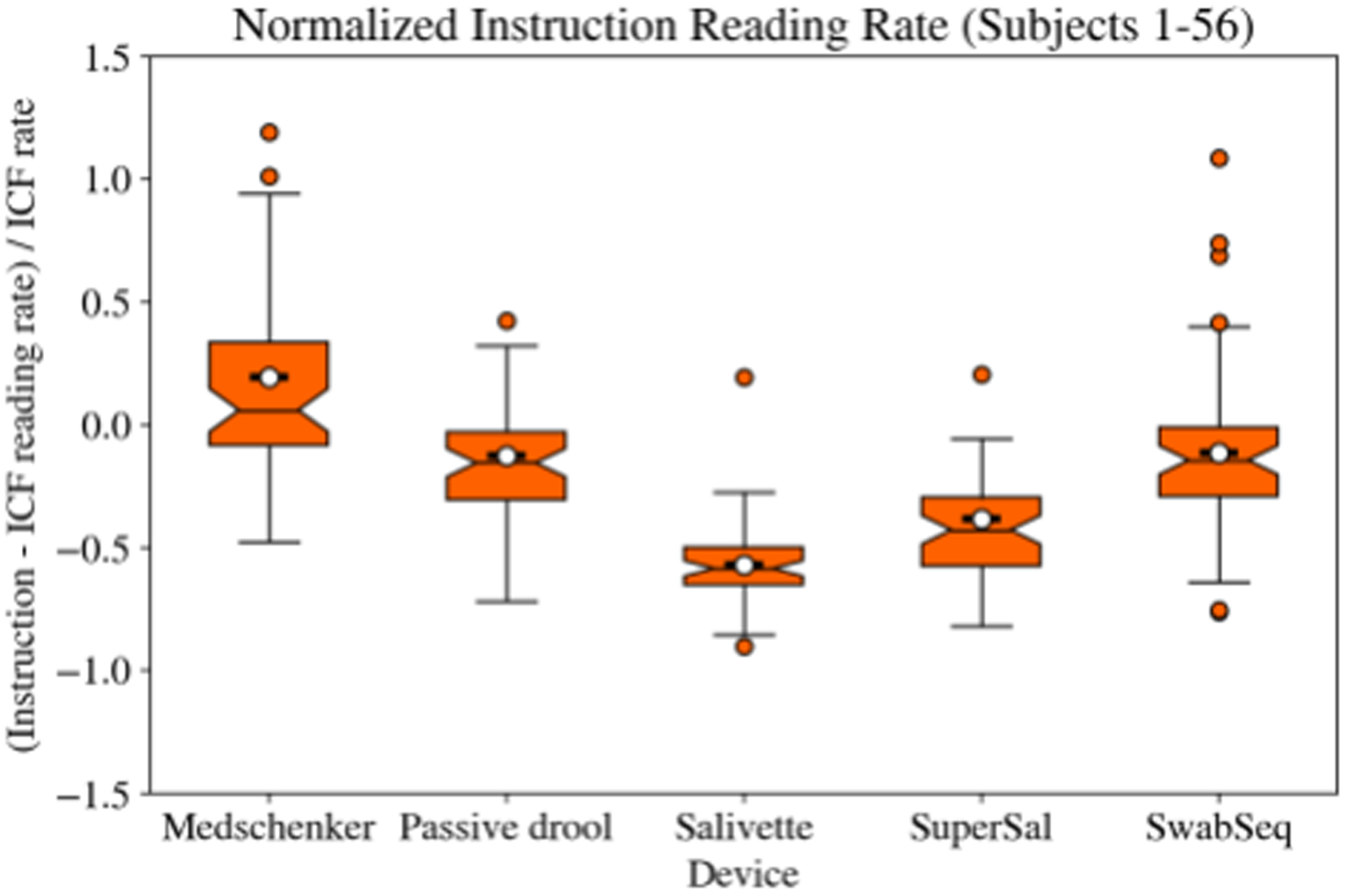
Instruction reading rate of each device, normalized by individual reading rate.

**Table 3.**
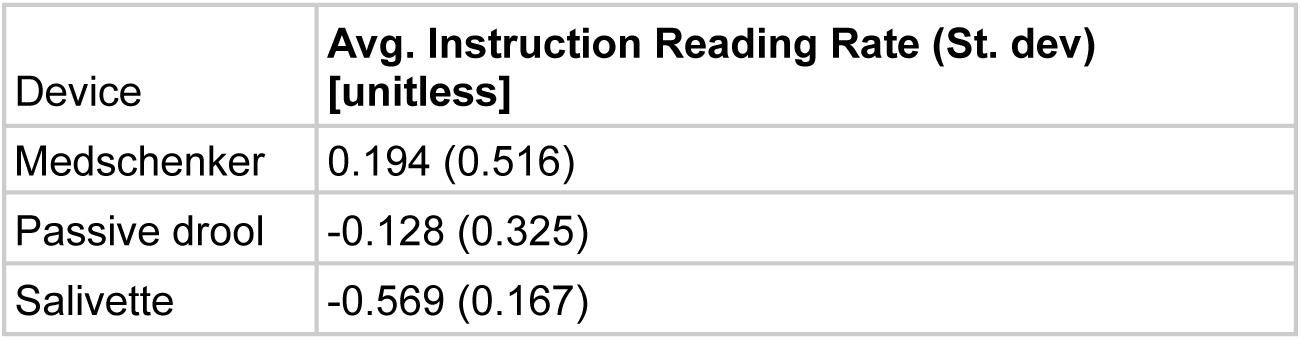
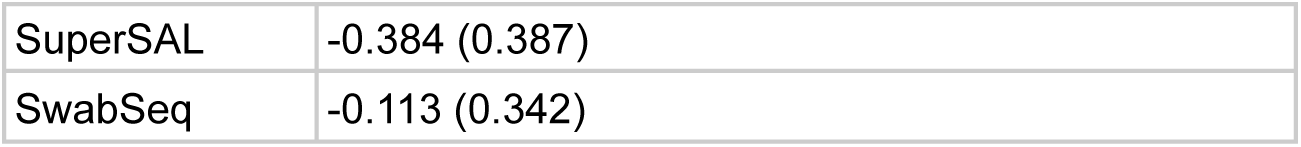
Average normalized instruction rate (Standard deviation)

| Device | Avg. Instruction Reading Rate (St. dev)<br>[unitless] |
| --- | --- |
| Medschenker | 0.194 (0.516) |
| Passive drool | -0.128 (0.325) |
| Salivette | -0.569 (0.167) |
| SuperSAL | -0.384 (0.387) |
| SwabSeq | -0.113 (0.342) |

The ranks in average normalized instruction reading rates were: Medschenker, SwabSeq, Passive drool, SuperSAL, and Salivette, from highest to lowest. Medschenker, which had the highest normalized rate, had a rate greater than zero, which suggests that the manual was easier to read or more quickly readable than a plain text-only document without jargon, such as the consent form that was used to measure the individual reading rates. However, it also had the widest interquartile range and the highest standard deviation, which implies that many of the users had a hard time reading the manual. The other devices all had rates below zero, even though the manuals had diagrams to supplement the text.

#### Leakage of saliva

All subjects were asked to answer in the survey whether saliva leaked onto their hands or clothes. Responses were given as “Yes” or “No,” and were translated into “True” and “False,” respectively, for data analysis. Results are shown in **Figure 4** and **Table 4**.

**Figure 4.**
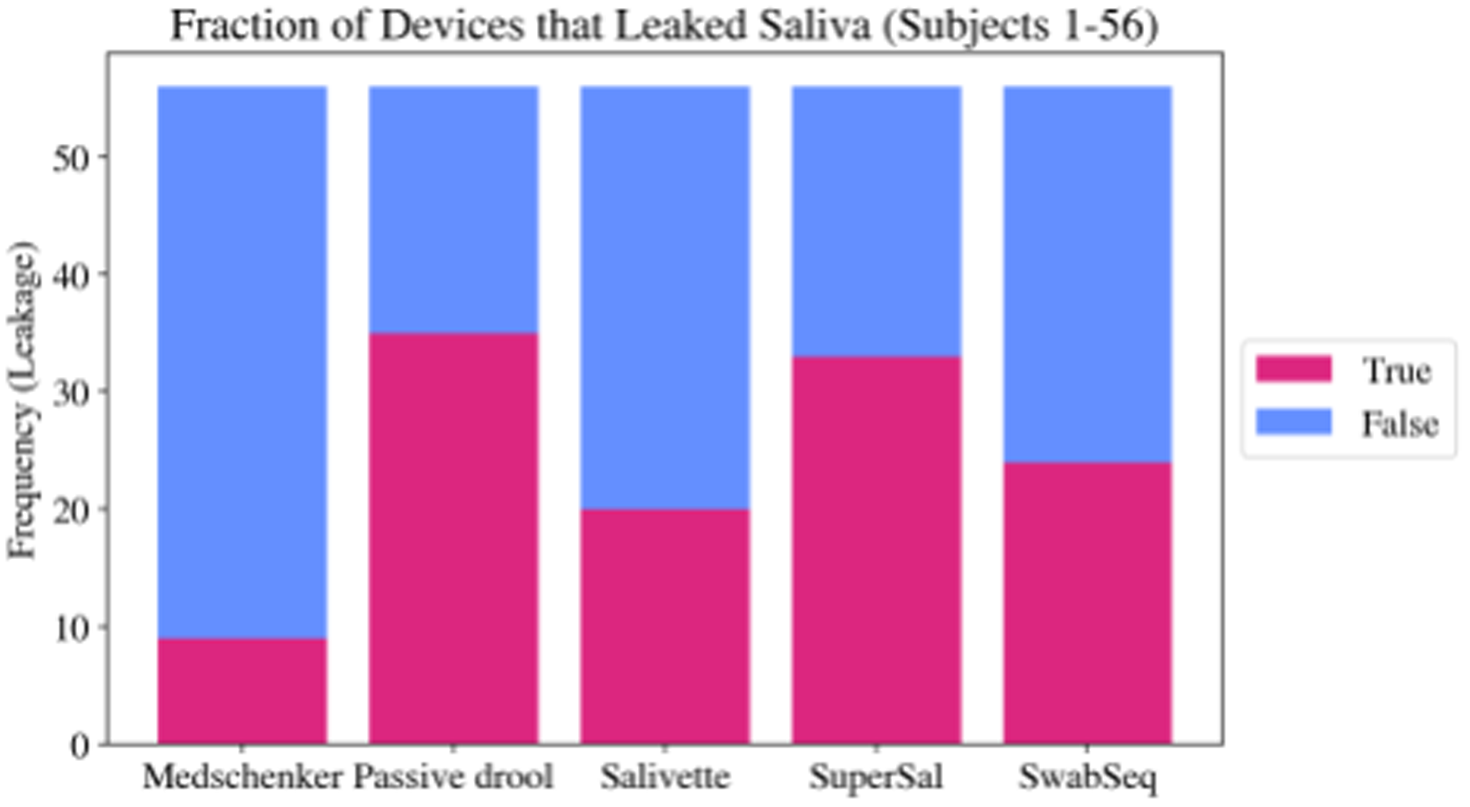
Fraction of devices that leaked saliva.

**Table 4.** Number and percentage of devices that leaked saliva.

| Device | Number of devices that leaked saliva, out of 56 | Percentage of devices with leakage (%) |
| --- | --- | --- |
| Medschenker | 9 | 16.1 |
| Passive drool | 35 | 62.5 |
| Salivette | 20 | 35.7 |
| SuperSAL | 33 | 58.9 |
| SwabSeq | 24 | 42.9 |

The fraction of devices that leaked were ranked as follows, from highest to lowest: Passive drool, SuperSAL, SwabSeq, Salivette, and Medschenker. Passive drool was closely followed by SuperSAL, and both devices had more than half of the devices resulting in leakage. Medschenker’s fraction of devices that leaked was significantly lower than those of the other devices.

#### Survey results

Subjects were asked questions on how difficult the instructions, assembly, and saliva collection for each device were. The answer choices presented were “Very easy”, “Easy”, “Fair”, “Difficult”, and “Very difficult”. Subjects could optionally write comments on each section. Results are shown in Figure 5, divided into three sections: instruction difficulty (Figure 5a), assembly difficulty (Figure 5b), and saliva collection difficulty (Figure 5c).

**Figure 5.**
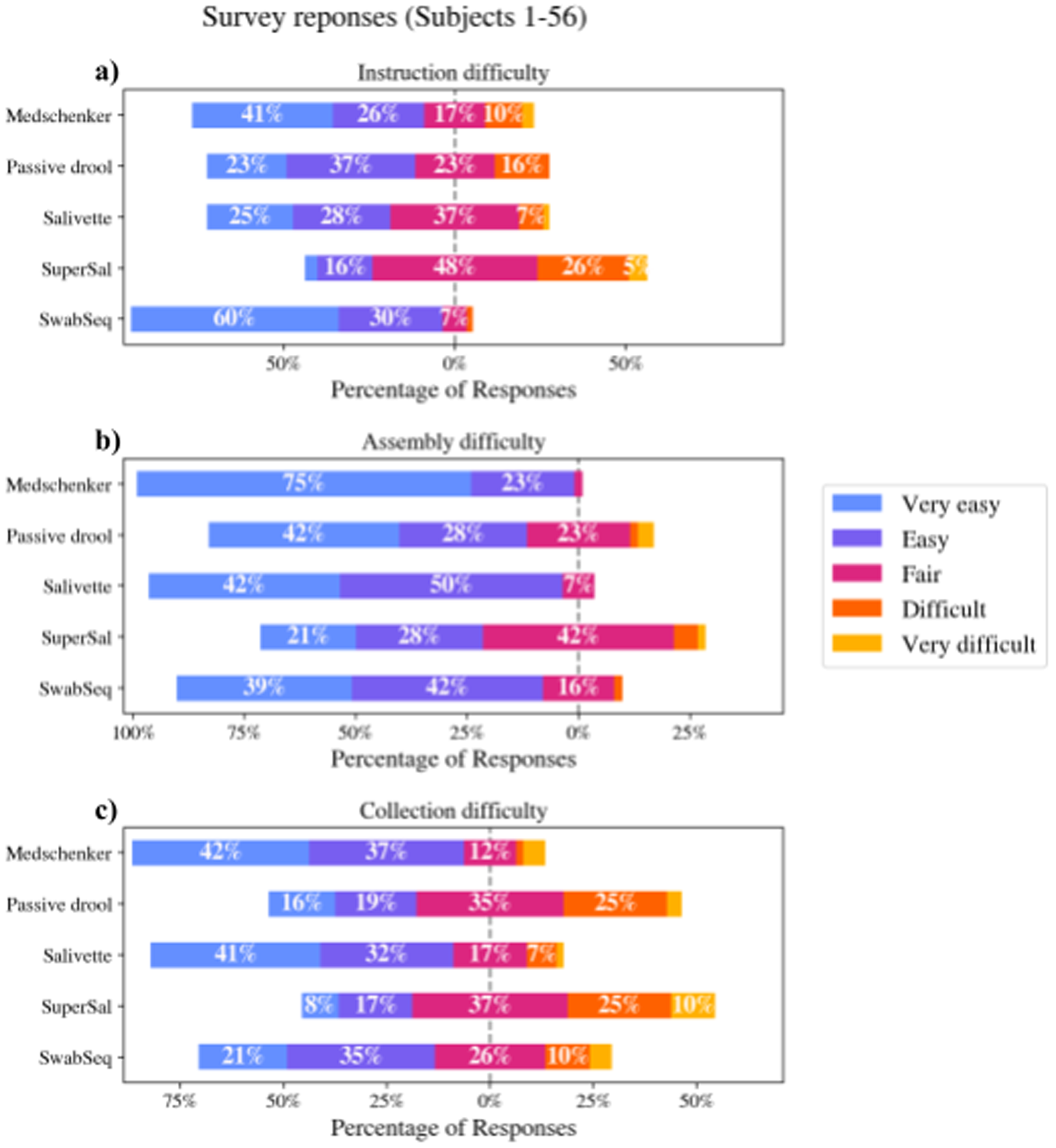
Survey responses on a) instruction difficulty, b) assembly difficulty, and c) saliva collection difficulty for each device.

The survey results showed that users found SwabSeq’s instructions the easiest to read (Figure 5a). Medschenker, Passive drool, and Salivette’s instructions were on average easy to read. Users found SuperSAL’s instructions the hardest to read, and the average difficulty of instructions for this device was “Difficult.”

The average assembly difficulty was easy for all devices (Figure 5b). Medschenker had the easiest assembly, closely followed by Salivette. While users found all devices fairly easy to assemble, SuperSAL had the highest number of “Difficult” and “Very difficult” responses.

Among the five devices, users found Medschenker and Salivette easiest to collect saliva with–both devices had over 70% of responses in either “Very easy” or “Easy.” (Figure 5c) Users found SwabSeq fairly easy to collect saliva with, but there were more responses in either “Difficult” or “Very difficult” for SwabSeq compared to Medschenker and Salivette. Passive drool had a slightly higher portion of “Very easy” and “Easy” responses than “Very difficult” and “Difficult” responses, and the user responses were more spread out between easy and difficult ratings than they were in other devices. Users found SuperSAL the most difficult device to collect saliva with, with an average difficulty of “Difficult.”

### Saliva flow rates

The purpose of this part of the study was to measure the increase in saliva flow rates to gauge the feasibility of collecting stimulated saliva with commercial saliva collection devices. Saliva flow rates by mass were measured in three conditions: unstimulated, water-stimulated, and citric-acid-stimulated saliva. The results are shown in Figures 6, 7, and 8.

**Figure 6.**
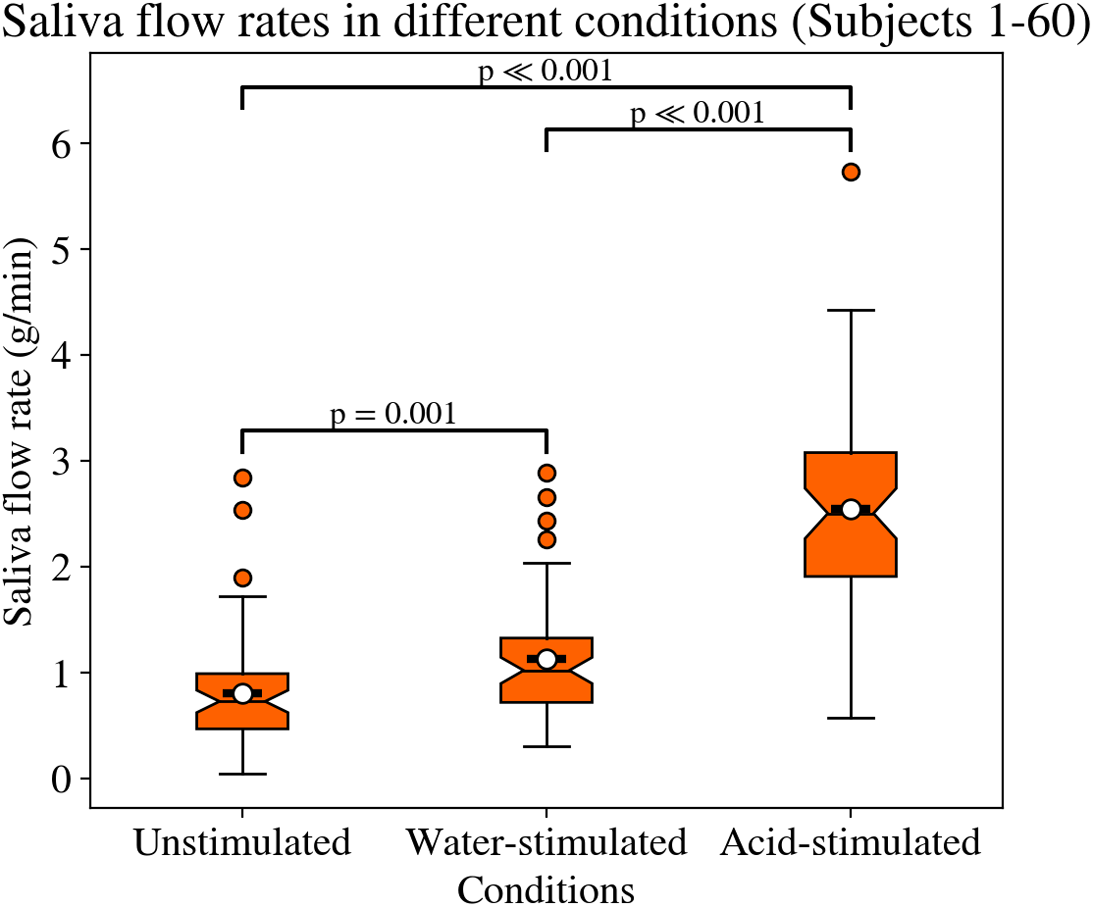
Saliva flow rates in unstimulated, water-stimulated, and acid-stimulated conditions for all subjects.

We observed statistically significant increases in saliva flow rate in water-stimulated and citric-acid-stimulated conditions compared to unstimulated conditions (Figure 6). The average citric-acid-stimulated saliva flow rate was around 1.74 g/mL higher than unstimulated saliva flow rate and 1.41 g/mL higher than water-stimulated saliva flow rates. In terms of fold change, the average acid-stimulated saliva flow rate was 3.17 times higher than the average unstimulated flow rate and 2.25 times higher than the average water-stimulated flow rate. The average water-stimulated saliva flow rate was around 0.33 g/mL higher than the average unstimulated flow rate, which translates to a 1.41-fold increase.

Furthermore, when the subjects were split by sex assigned at birth, we observed significant differences between male and female saliva flow rates for all three conditions (Figure 7, Table 5). For unstimulated salivation, male saliva flow rates were 0.279 g/min higher, or 1.42 times higher than female saliva flow rates. For water-stimulated saliva, male saliva flow rates were 0.428 g/min higher, or 1.41 times higher than those of female subjects. For acid-stimulated saliva, the difference was even greater than that in water-stimulated saliva: male salivation flow rates were 0.690 g/min higher, or 1.28 times higher than those of female subjects.

**Figure 7.**
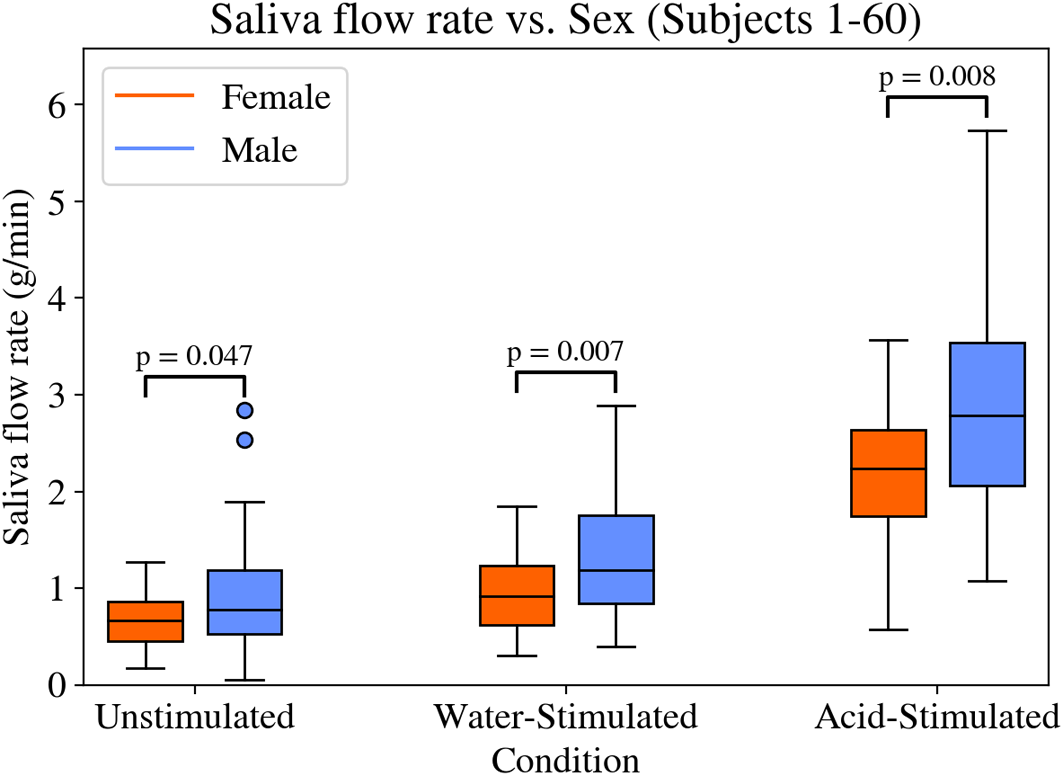
Saliva flow rates in different conditions, separated by sex of subjects.

**Table 5.**
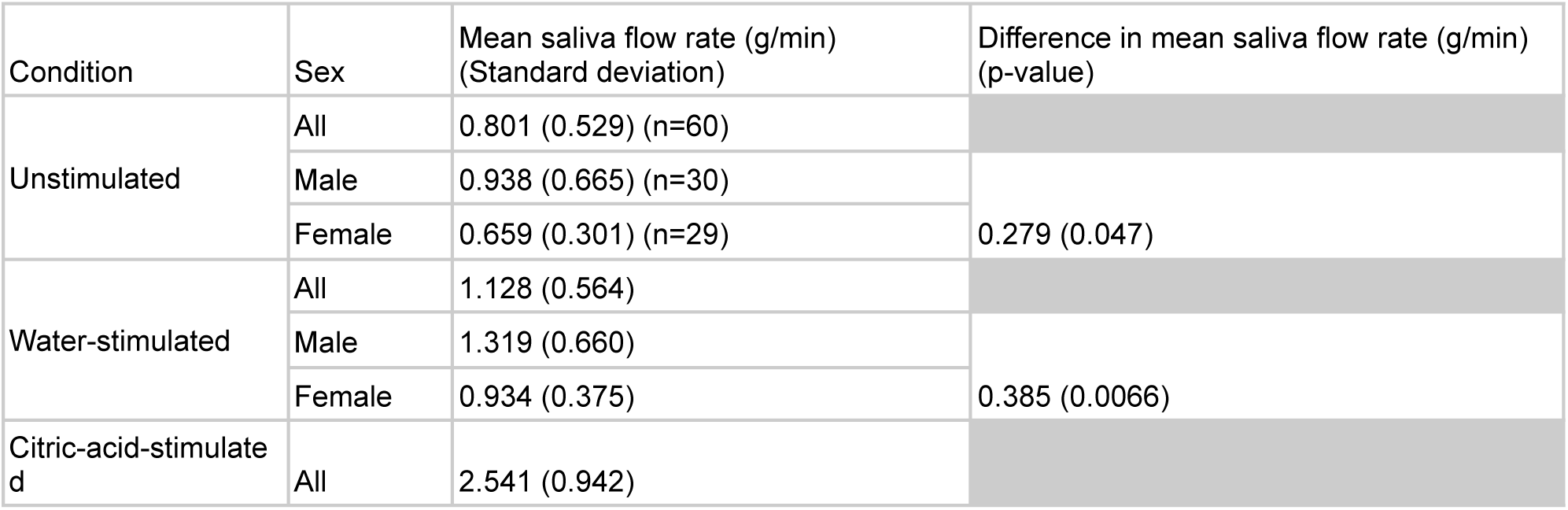
Average saliva flow rate (g/min) (standard deviation)

In the samples collected over time, the unstimulated saliva flow rates did not change significantly (Figure 8). Water-stimulated saliva flow rates dropped significantly between 30 seconds and 60 seconds: the difference in mass of collected saliva dropped by 0.23 grams between the 30 second and 60 second replicate, which translates to around 32% decrease in mass. The difference between the 60 second and 90 second samples was not significant. For citric-acid-stimulated saliva, significant decreases in saliva flow rates were observed between all three replicates. From the 30 second to 60 second samples, the collected saliva mass dropped by 1.02 grams, which is equivalent to a 51% decrease. From the 60 second to 90 second samples, the collected saliva mass dropped by 0.21 grams, which translates to a 21% decrease in mass. These results again show that water as a stimulus has an effect that lasts up to around 60 seconds after stimulus introduction, whereas citric acid has a longer-lasting effect that is observed even at 90 seconds after stimulus introduction.

**Figure 8.**
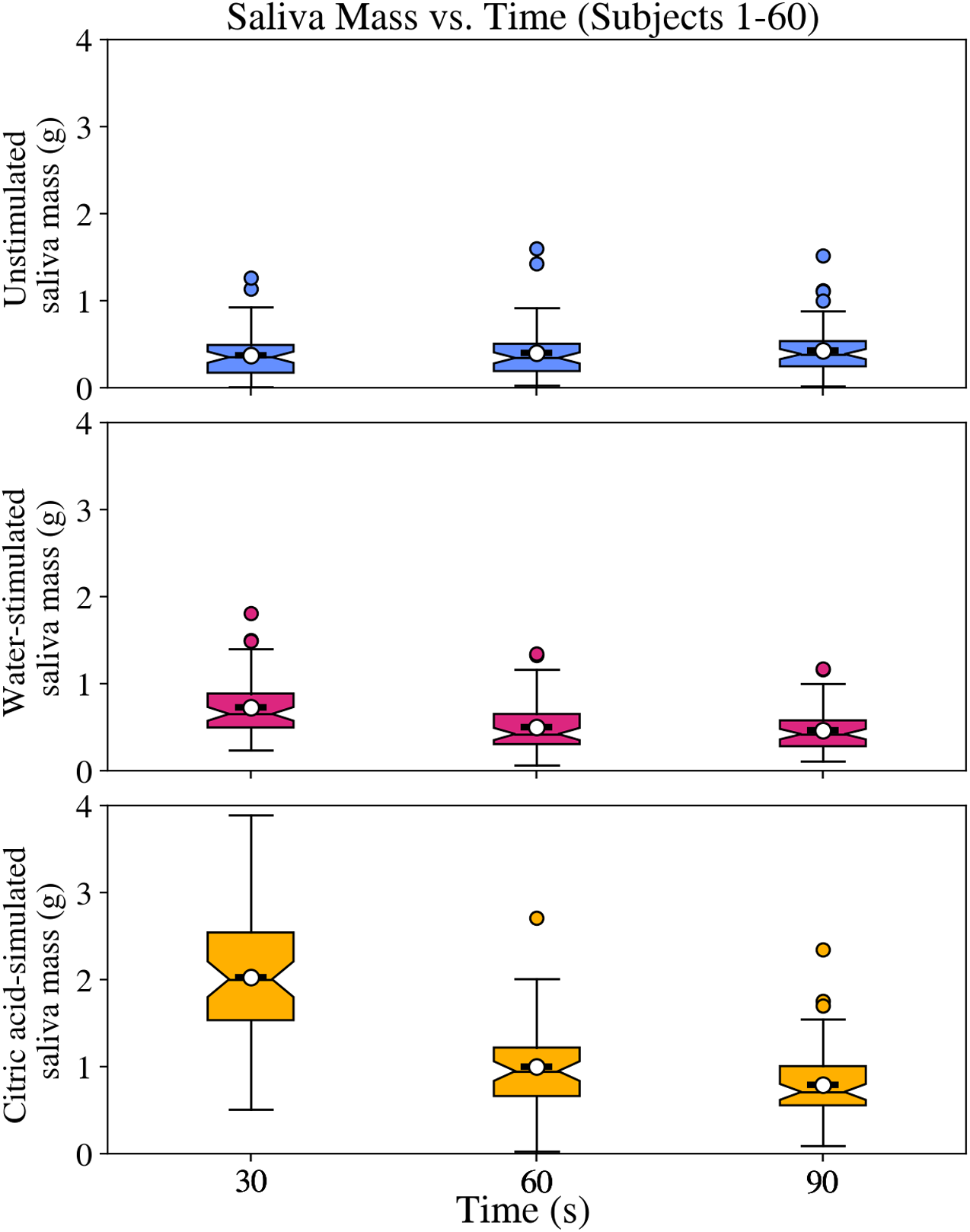
Change in saliva mass over time after stimulus introduction. Stimulus was introduced at t=0, and three samples were collected for each condition, in 30-second intervals.

## Conclusions

In the results, we summarize the major outcomes for each metric we measured in our study, as well as how each device performed with respect to those metrics. With respect to just the quantitative and qualitative metrics such as the saliva collection rate, instruction reading rate, and presence of leakage, Medschenker’s saliva collection kit performed well on most metrics, while SuperSAL did not perform as well. However, even the Medschenker device did not satisfy some aspects of usability, as can be seen in the survey responses for the device. This shows that a device that satisfies all metrics–a device that allows for efficient saliva collection, is easy to use, and collects saliva without leakage–would be ideal for saliva collection. The design and distribution of such devices could greatly help standardize saliva collection, as there are a myriad of saliva collection devices in the market that are suited for specific applications or aspects of saliva collection, and this overwhelming variety in saliva collection devices contributes to the unstandardized nature of current saliva collection protocols.

### Remarks on each device

Below are additional remarks specific to each device, following analysis of the device’s design and comments left by the subjects on the survey.

#### Medschenker

Medschenker yielded efficient saliva collection, as can be observed in the normalized collection rate–and the highest normalized instruction reading rate out of the five devices (Figures 2, 3, Tables 2, 3). Medschenker also yielded the least leakage out of the five devices (Figure 4, Table 4). On average, users found Medschenker’s instructions easy to understand and the device easy to collect saliva with (Figure 5). However, comments left on the survey suggested that the instructions were too long, contained extraneous information directed at the sample handlers and not the users, and were ambiguous on how much volume was supposed to be collected, as the collection tube lacks any volume graduations (Figure 9b, 9c). Comments on saliva collection stated that the big funnel allowed for easy saliva collection, but that the volume of saliva to collect is too large (Figure 9b, 9c).

**Figure 9.**
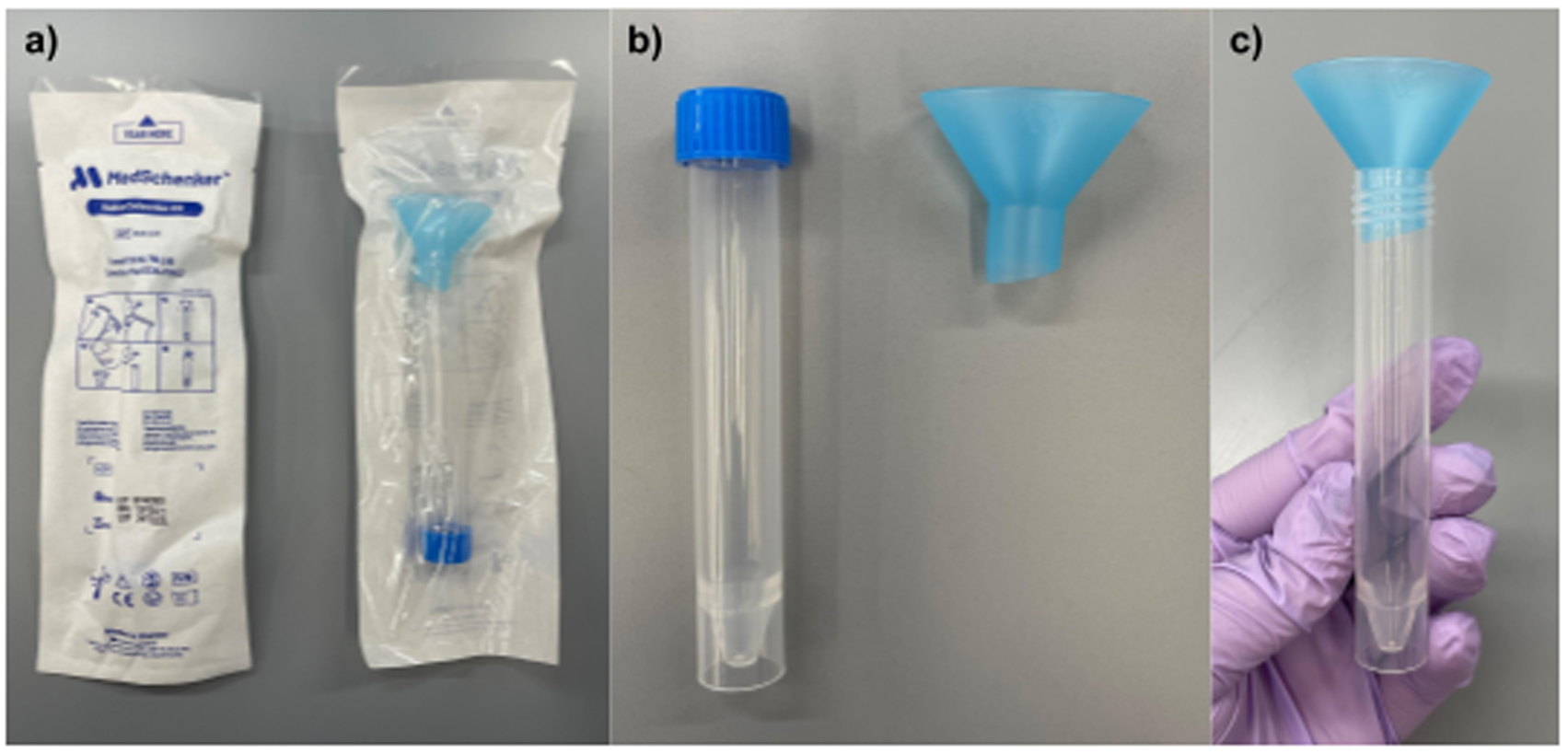
The Medschenker saliva collection device. a) The device in its packaging, b) the device taken out of the packaging, and c) the set-up of the device for saliva collection.

#### Passive drool

Passive drool was third in both normalized saliva collection rate and normalized instruction reading rate (Figures 2, 3, Tables 2, 3). On the survey, Passive drool had mixed opinions on the difficulty of saliva collection but its instructions were rated easy to read on average (Figure 5). However, comments on the survey stated that the instructions contained some confusing diagrams and jargon, and that it was difficult to check how much saliva was collected. Passive drool’s major drawback was that it had the most leakage out of the five devices, with over half of the devices resulting in leakage (Figure 4, Table 4). Through the comments left on the survey and our own analysis on the design of the device, we identified two sources of saliva leakage: the narrow saliva inlet, and the wide air vents in the saliva collection aid (SCA) (Figure 10c). As many users stated through the comments, the narrow straw-like saliva inlet made it difficult for users to guide the pooled saliva through the SCA into the collection tube. However, many more comments stated that saliva leakage came from the gaps between the SCA and the collection tube, which was intentionally built as an air vent for the device. When liquid enters a container, there must be a way for air to get displaced as the liquid takes the place that was previously held by the air. If the air vent is too wide, even a small pressure exerted, such as the user blowing air through the SCA while collecting saliva, saliva can exit through the air vent along with the air, causing leakage. As there were four such air vents between the SCA and the collection tube, leakage of saliva happened quite frequently as users either exhaled into the SCA or simply collected too much saliva that ended up flowing over the tube.

**Figure 10.**
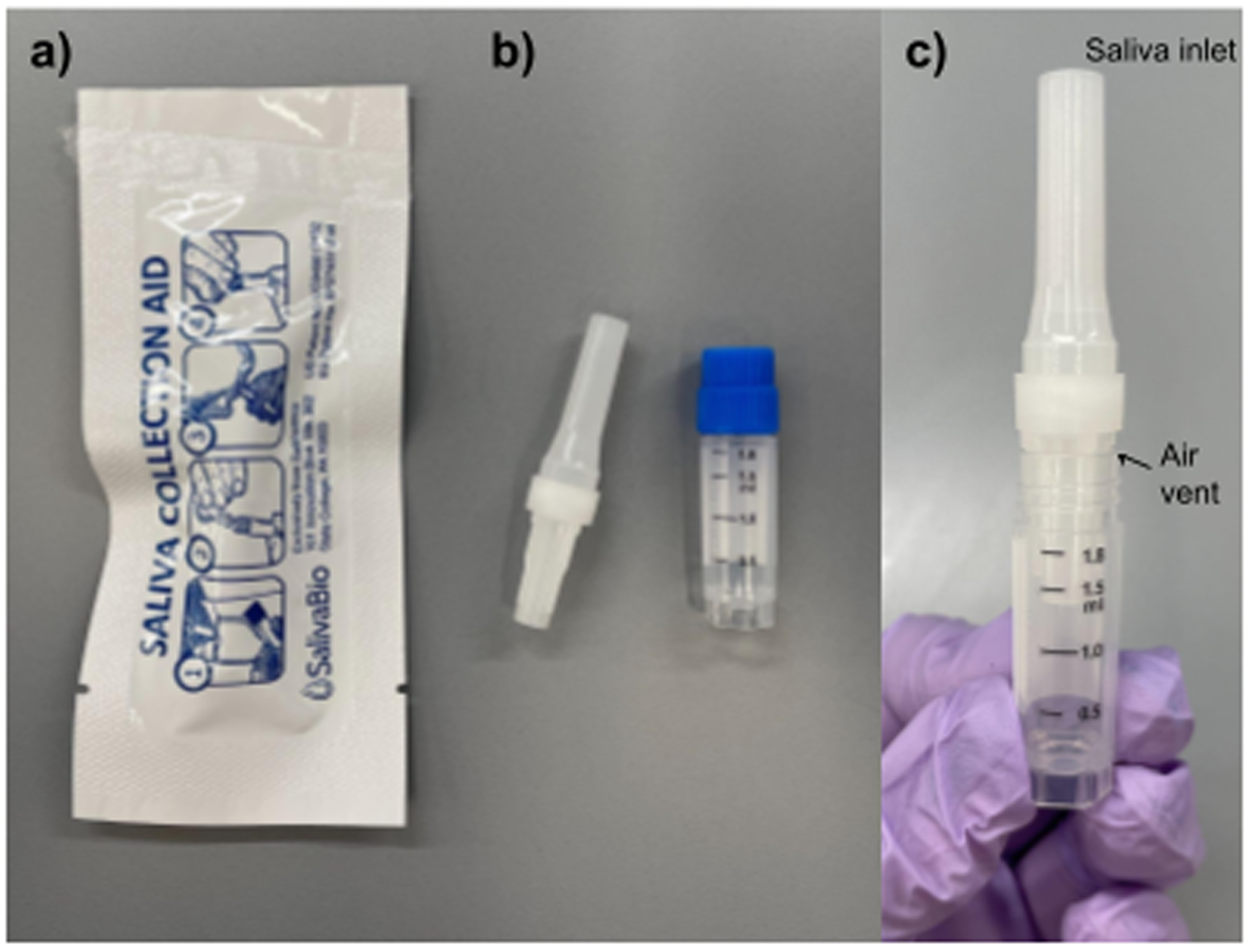
The Passive Drool saliva collection device. a) The device in its packaging, b) the device taken out of the packaging, and c) the set-up of the device for saliva collection, with the saliva inlet and air vent marked.

#### Salivette

Salivette had the highest normalized saliva collection rate out of the five devices, making it the most efficient device at saliva collection (Figure 2, Table 2). It also had the second lowest leakage rate, most of which we identified to have come from the act of touching the saliva-soaked swab when the users placed the swab back into the tube after collection (Figure 4, Table 4). However, it had the lowest normalized instruction reading rate, which implies that users had a hard time reading and understanding the instructions (Figure 3, Table 3). Even though the normalized instruction reading rate was the lowest, the survey results for the instruction difficulty were rated easy on average (Figure 5). Similarly, the assembly and saliva collection with Salivette were rated easy on average on the survey (Figure 5). However, comments on the survey showed additional concerns with this device. Many users stated that the font was too small for the instructions, which was understandable as the instruction manual had instructions written in more than 20 languages all on the same page, thus constraining the space for text. Also, users stated that the diagrams on the instructions were ambiguous and strange. Many users also stated that they were confused about the inner tube (Figure 11b), which exists to hold the swab in place while the collection tube is centrifuged after collection to extract saliva. Also, many users complained that it was unclear how much saliva was collected and that they were concerned about touching the saliva-soaked swab with their hands when they returned the swab into the tube after collection (Figure 11b). There were mixed opinions on the swab, with many users stating that the swab made it easy to collect saliva because they did not have to spit, and even more users stating that the swab was unpleasant in their mouths. Although this point was not stated in our results as this study was focused on the perspective of the user, we believe the fact that the collection tube needs to get centrifuged in order to extract saliva may affect its usability in point-of-care settings, as it requires facilities or the users to have access to a centrifuge.

**Figure 11.**
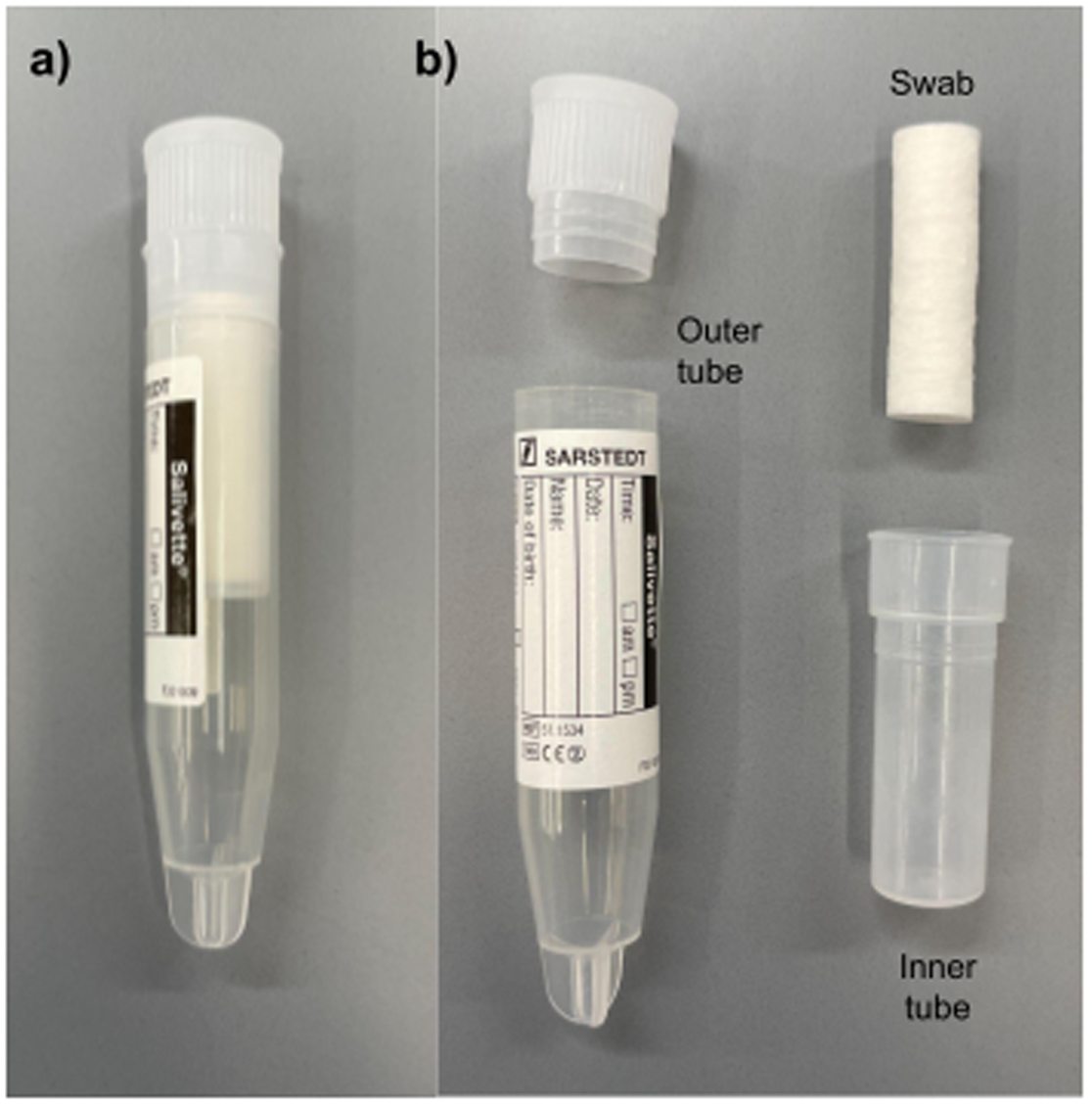
The Salivette saliva collection device. a) The device does not have external packaging and is presented this way, and b) the anatomy of the device. The set-up of the device for saliva collection was not pictured, as the saliva collection process simply requires the user to place the swab (labeled) in the mouth for 2 minutes and then to place it in the inner tube (labeled) that goes inside the outer tube (labeled).

#### SuperSAL

SuperSAL had the lowest normalized saliva collection rate and normalized instruction reading rate out of the five devices, making it the least efficient and most difficult device to use with respect to quantitative metrics (Figures 2, 3, Tables 2, 3). Their survey responses showed that out of the five devices, users found SuperSAL’s instructions, assembly, and saliva collection the most difficult. In the comments, many users stated that they were confused about the instruction through comments, and many of the users that commented specifically stated that the presentation of the compression tube and the absorbent swab was confusing, as they were combined inside the packaging while the instructions depicted them as separate components (Figure 12a). Some comments also pointed out that technical terms such as “Eppendorf tubes’’ should not have been included in the instructions. Most comments on saliva collection with SuperSAL stated that it took too long to collect saliva with the device. Many users also complained that the swab was unpleasant and uncomfortable, and that it was tedious to keep checking the color change of the swab’s Sample Volume Adequacy Indicator (SVAI), which is a dot on the swab that changes color once enough saliva has been collected (Figure 12b). Some users also stated that it was uncomfortable and even painful when compressing the swab to extract saliva. SuperSAL had the second highest leakage rate, with over half of the devices resulting in leakage (Figure 4, Table 4). Through the comments, we identified the source of leakage as the gap between the compression tube and the collection tube, due to the foaming of saliva as it gets aspirated during the compression of the swab. Also, our analysis showed that the volume of saliva collected with SuperSAL was inconsistent, as users could only rely on the color change of the SVAI during collection; there were even cases in which the SVAI changed color but there was no saliva extracted.

**Figure 12.**
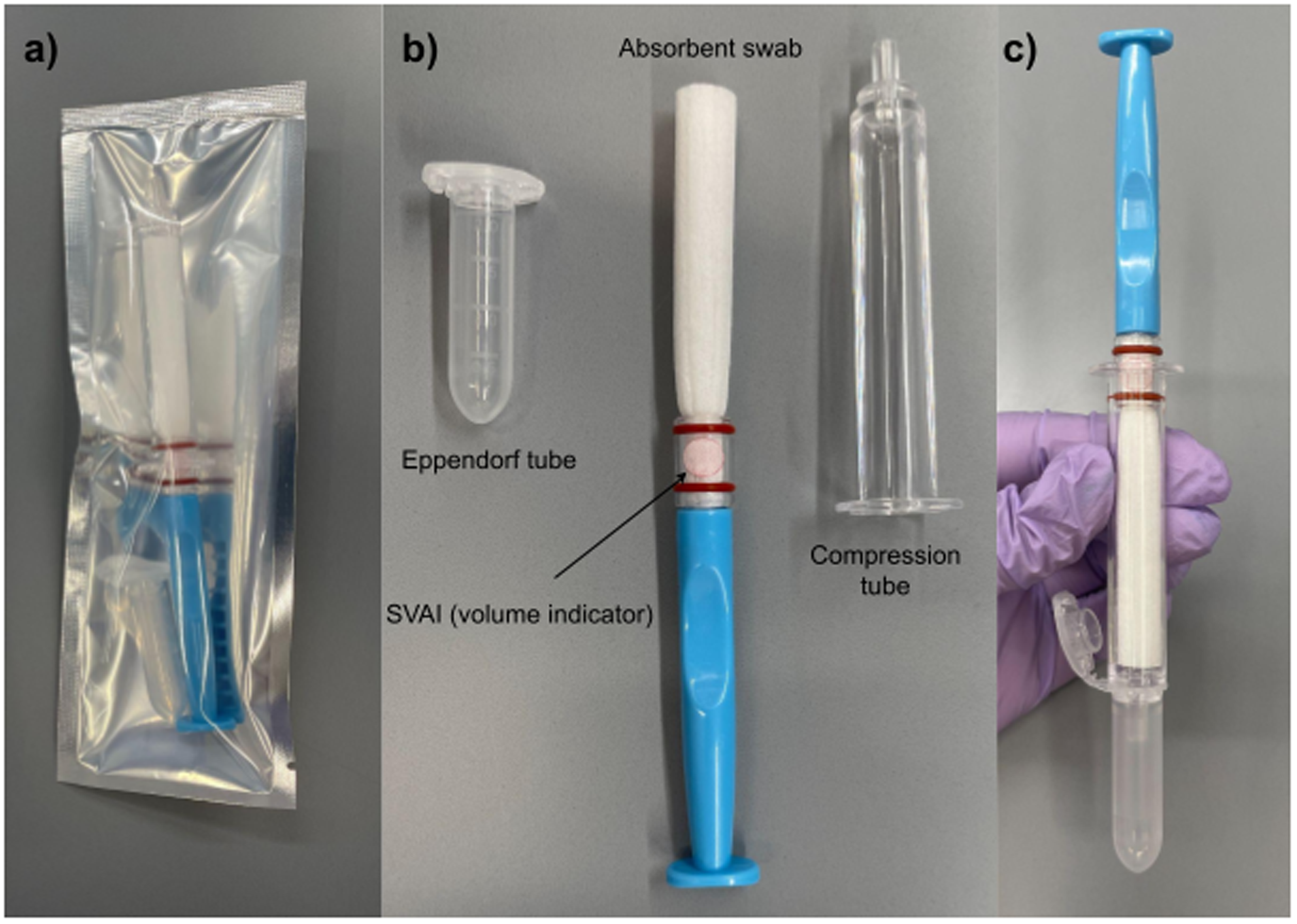
The SuperSAL saliva collection device. a) The device in its packaging, b) the device taken out of the packaging and disassembled, and c) the set-up of the device for saliva extraction after saliva collection.

#### SwabSeq

**Figure 13.**
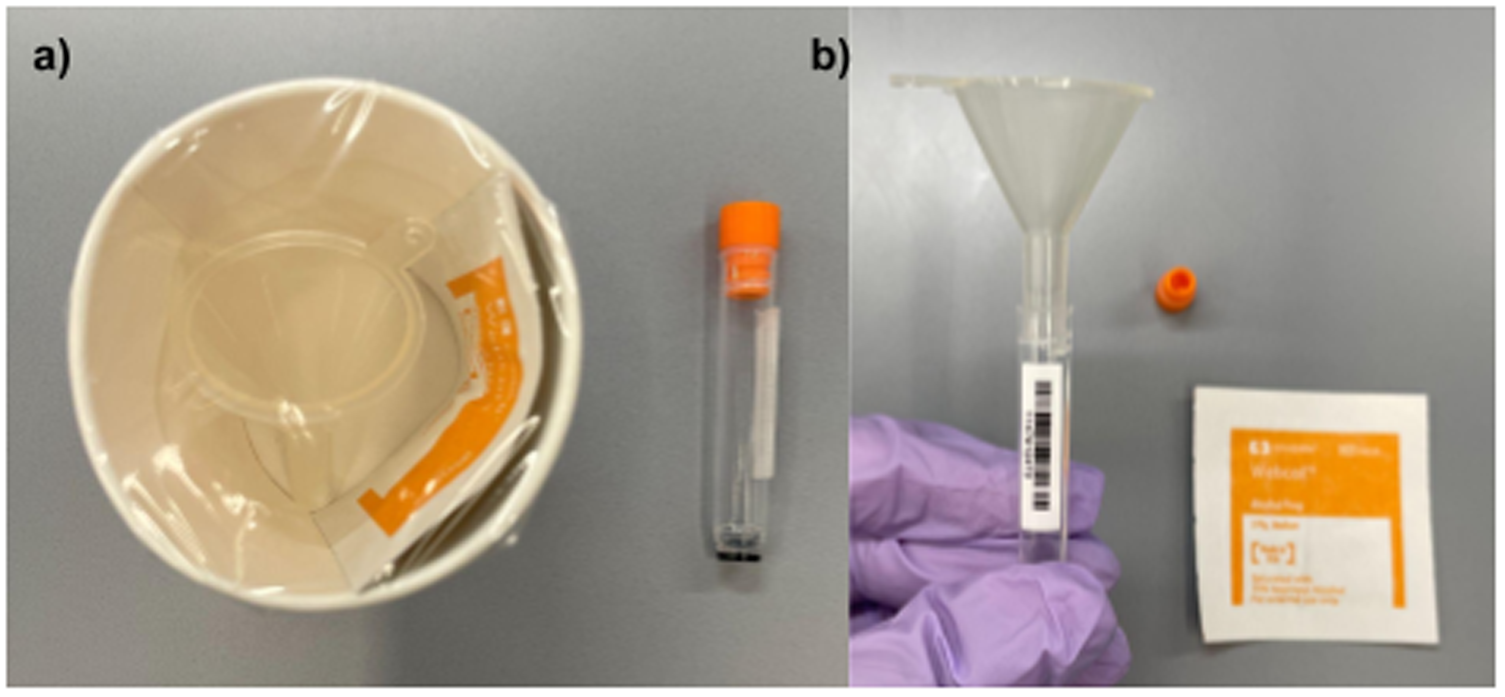
A cryovial and funnel saliva collection kit, referred to as SwabSeq in this study^†^. a) The packaging of the device as was presented during the surveillance tests on Caltech campus, and b) set-up of the cryovial and funnel for saliva collection, with the cryovial lid and alcohol swab in the background.

SwabSeq’s funnel and cryovial set-up had the second lowest normalized saliva collection rate out of the five devices, and the second highest normalized instruction reading rates (Figures 2, 3, Tables 2, 3). This was reflected in the survey responses on the difficulty of instructions, as SwabSeq was rated as the device with the easiest instructions. In the comments, users stated that SwabSeq’s instructions on “[thinking] about [their] favorite food” were helpful in producing more saliva. On the other hand, even though the survey response for SwabSeq’s saliva collection difficulty was rated easy on average, comments on the survey helped explain the low normalized saliva collection rate. Many users commented that saliva got stuck in the funnel, and that there was leakage and foaming of saliva. The tight fit of the narrow funnel in the mouth of the collection tube did not allow air to leave the collection tube as the viscous saliva plugged the funnel. This resulted in saliva getting stuck in the narrow part of the funnel before it could enter the collection tube, and users had to lift the funnel from the collection tube to let saliva through, which in turn resulted in leakage of saliva. Both Passive drool and SwabSeq had high leakage rates due to the problem of air venting of the device, but for opposite reasons: Passive drool had leakage because the air vents were too large, and SwabSeq had leakage due to the lack of an air vent.

### Choosing a saliva collection device

Among the metrics we measured for the five devices we chose were saliva collection efficiency, usability (ease of use), and presence of leakage. Below we list characteristics of saliva collection devices that an ideal device should have.

#### Performance

An ideal device should collect saliva efficiently and effectively. An efficient saliva collection device will be useful in a situation where fast throughput of tests is important, such as a mass-testing scenario where multiple people are collecting saliva inside a testing tent. The device should also allow collection of sufficient saliva that is needed for the downstream processes for the saliva sample.

#### Usability

The device should be easy to use for the user in all aspects: the instructions should be easy to read and comprehend, the assembly should be simple and as minimal as possible, and the saliva collection with the device should be minimally invasive and easy to do. The instructions should be concise, but detailed enough to fully describe everything the user needs to know from the assembly and use of the device to how much saliva should be collected. Instructions should be as free of technical terminology as possible, and should ideally contain helpful diagrams to visualize some processes. The assembly of the device should be intuitive in that the device is presented in the packaging in a way such that the user can easily guess what each part is for and how they are to be assembled together. Ideally, the device should be designed robustly such that the user cannot easily make mistakes assembling the device. Saliva collection with the device should be minimally invasive and cause as little discomfort for users as possible. Furthermore, the user should be able to easily perceive if they have collected a sufficient amount of saliva, whether it be through visually inspecting the level of saliva collected or through some other kind of indication such as a moisture indicator that turns color when enough saliva has collected, like the SVAI for SuperSAL. Lastly, the device should only collect enough saliva that is necessary for the assay the saliva is to be used for, as it is difficult for users to readily produce large amounts of saliva.

#### Hygiene

The device should collect saliva in a manner that minimizes saliva leakage. Saliva leakage, if the outer sides of the device or the user’s hands are not sterilized correctly after, can be a source of secondary infections if the saliva contains pathogens. Furthermore, saliva leakage can be a source of discomfort for the users and sample handlers receiving the saliva samples. Ideally, the device’s design, especially with respect to the air venting mechanism of the device, should account for the foaminess and viscosity of saliva and minimize the potential sources of saliva leakage. Furthermore, user instructions should indicate correct ways of collecting saliva that can minimize leakage, and explain methods of sanitizing the device and the user’s hands if there is saliva leakage.

#### Saliva viability

The saliva collection device should collect saliva that is viable for downstream purposes. This can mean both quantity and quality of saliva: enough volume of saliva should be collected, and the collected saliva should not be contaminated or altered in a way that can compromise the biomarkers of interest. An ideal device should collect clear, whole saliva that is not too viscous and does not contain sputum, food debris, or chemicals.

#### Cost and logistics

The saliva collection device should be inexpensive and easy to source. An ideal device would have components that are easy to source and can easily be replaced with an alternative in the case of supply chain issues. Also, the saliva collected should require as little post-processing as possible, especially if the saliva is to be collected and analyzed in a point-of-care setting with limited personnel and resources. Furthermore, the device should avoid the requirement of specific reagents, machinery, or protocols to process the saliva samples, as these aspects can all affect the accessibility of the device.

## Discussion

### Measurement precision

Measurement precision issues in volume measurements of saliva samples were observed in the part of the study where individual saliva flow rates were measured. To mitigate this issue, we used the mass of saliva to calculate saliva flow rates instead of volume. In order to convert the mass of saliva collected with Salivette to volume such that the collection efficiency of Salivette could be compared to those of other devices, the median density value of unstimulated saliva from prior literature (1.007 g/mL) was used.

### Population of subjects for study

The population for this study consisted of healthy adults between the age of 18 and 65. While this study may have shown preferences and performances of users in this age group, the results should not be extended to populations outside of this age group. When choosing saliva collection methods for older adults, users with disabilities, and pediatric patients, the accessibility or usability of the device should be considered, as well as the natural decrease in saliva flow rate with age (9). For instance, the Salivette swab may not be suitable for those who cannot keep the swab in their mouths, as the swab may pose choking hazards if accidentally swallowed. Devices that require the use of two hands, such as SuperSAL when the user compresses the swab to extract saliva, may not be suitable for those with limited mobility or strength in their hands. Future research should focus on determining the most appropriate method of collecting saliva for demographics outside the 18-65 age group.

### Stimulated saliva collection

In this study, subjects were asked to collect unstimulated saliva with the given saliva collection devices. Many comments on the survey stated that it was difficult to produce saliva while using the devices. Although real-life situations for saliva collection would not usually require patients to collect sample after sample as subjects were asked to in this study, it is worth mentioning that helping people produce saliva more readily may pose many benefits, as long as the method of increasing saliva production does not compromise the viability of the sample. In the perspective of efficiency and practicality, increased saliva flow rate can be translated to increased efficiency in saliva collection, which may result in higher throughput of saliva collection in a mass-testing scenario, for instance. Related to the previous point in discussion about the accessibility of saliva collection devices, patients who cannot readily produce saliva, such as patients with xerostomia (dry mouth) or older adults who naturally have lower saliva production, may benefit from increased saliva production during saliva collection. In our study, we measured the unstimulated saliva flow rates as well as stimulated saliva flow rates, and showed that stimuli such as citric acid solution can significantly increase saliva flow rates. While citric acid is one of the strongest known gustatory stimuli for increasing saliva flow rates (10), it may alter the pH of saliva samples and render the sample unusable for certain assays. Although we did not measure the pH of the collected stimulated saliva samples in this study, future research could explore the use of citric acid as a gustatory stimulus for temporarily increasing saliva flow rates, and find the optimal concentration of citric acid that can increase saliva flow rates to aid saliva collection while keeping the pH of saliva within reasonable levels.

## Methods

### 1. Aim, design, and setting of the study + study registration

The aim of the study was to measure performances of saliva collection devices, and individual saliva flow rates under stimulated and unstimulated conditions for normalization. The study was deemed exempt by the Caltech IRB due to the minimal-risk nature of the tasks involved. The number of subjects to be tested was determined by statistically estimating the needed sample size as follows.

The equation for sample size *n* is given by

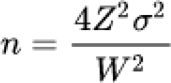

where *Z* is the Z-score for a given confidence interval, *σ* is the standard deviation, and *W* is the width of the confidence interval. Based on prior literature states we assumed that the mean flow rate for unstimulated saliva was 0.58 mL/min with a standard deviation of 0.24 mL/min (11). With the assumption that saliva flow rates are distributed normally with a mean of 0.58 and standard deviation of 0.24, we simulated values of saliva flow rates by randomly generating samples from this normal distribution. With these values, we calculated the standard deviation of time needed to collect 1.5 mL as 1.97 minutes, and with these we determined the margin of error, W/2. The mean flow rate of unstimulated saliva in the same publication was given as 0.58 mL/min, which translated to 2.59 minutes to collect 1.5 mL of saliva. Setting the margin of error to be ±20% around this mean, we calculated that *W* = 1.032 minutes. The Z-score for a 95% confidence interval is given as 1.96 and the confidence interval width *W* is 1.032 minutes. Using the equation above, we determined that the minimum sample size *n* was 56, which we rounded up to 60. The actual number of subjects recruited was 60, but the last 4 subjects completed the study without the MedSchenker saliva collection device due to unforeseen supply issues. The study was registered after the analysis of the first 30 “exploratory” samples but before the opening of the last 30 “confirmatory” samples.

The study was registered on Open Science Framework (OSF) Registries [https://doi.org/10.17605/OSF.IO/EFAP6] after the analysis of the first 30 samples but prior to the access to and analysis of the latter 30 samples. Significant results from the analysis of the first 30 samples, which we refer to as “exploratory” samples, were pre-registered. Analysis results from the latter 30 samples, or “confirmatory” samples, were compared with the results from the exploratory samples.

### 2. Recruitment of subjects

Subjects were recruited by fliers, emails, and word-of-mouth advertising. All subjects were directed to the same QR code that links to a Google Forms that contains information about the study and a text entry for subjects to provide their email addresses for scheduling purposes. The subjects were contacted through email to schedule their appointment. Upon recruitment, subjects were asked for consent to fast for 30 minutes before the session in order to simulate a typical saliva collection protocol. During the 30 minutes, subjects were asked to avoid eating, drinking, smoking, chewing gum, or brushing their teeth. Only subjects above the age of 18 were recruited for the study.

### 3. Informed Consent

At the beginning of the session, subjects were asked to optionally present their age and assigned sex at birth. Then, subjects were asked to read the Informed Consent Form (ICF) while being timed. This time was used to calculate the subject’s base reading rate, in words per minute, to normalize the instruction manual reading times for each device. After reading the ICF, subjects were asked to check a box stating “Agree” or “Disagree.” The signed ICF was collected by the researcher with the rest of the data collected during the session, and copies of the signed ICF were given to the subject per request.

### 4. Saliva collection with devices

After obtaining informed consent from subjects, the researcher presented five saliva collection devices in random order to the subject. For the collection tube of the MedSchenker saliva collection kit, the cryovial in the SwabSeq collection kit, and the collection tube for SalivaBio’s passive drool device, the line for the recommended volume was marked with a custom-made 3D-printed tool, as the lines specifying the target volume were not clear. The mass of the unused Salivette device was measured at the beginning of the study, such that the mass of the collected saliva could be measured by weight.

For each device, subjects were asked to read the instruction manual from start to finish while being timed. The subjects were then asked to collect saliva with the device while being timed from when they start using the device until they finish collecting the specified amount of saliva. The volume of saliva collected for each device was given as follows: 1 mL with MedSchenker, 1 mL with Passive drool, and 0.5 mL with SwabSeq. For SuperSAL, saliva was collected until the volume indicator (SVAI) turned red in color. For Salivette, subjects were asked to keep the swab in their mouths for 2 minutes. Subjects were allowed to refer back to the manuals when they were confused on how to use the device. The researcher did not give instructions on or comment on the user’s behaviors unless the subject or the device was in danger. After using each device, subjects were asked to answer a questionnaire on their experience with the device, with questions about whether there was leakage of saliva, how difficult they found the assembly/instructions/saliva collection method of the device, with optional comments. Each survey was collected and sealed in an envelope until data analysis.

### 5. Saliva production rate measurement

After collecting saliva with all five devices, the subject’s individual saliva production rate was measured. Saliva was collected in three conditions: unstimulated, water-stimulated (control), and citric-acid-stimulated. The citric acid simulation was given in the form of a 0.02 M solution of food-grade anhydrous citric acid dissolved in distilled water. The water was given as a control for measuring the effects of having liquid in the mouth. For each condition, three replicates were collected in 30-second intervals. In the unstimulated case, the subject was first asked to remove all saliva from their mouth, pool saliva for 30 seconds, spit into a falcon tube with a funnel, and repeat for the next two replicates. For the water and citric-acid stimuli, subjects were asked to gently gargle a comfortable amount of the liquids in their mouths for 30 seconds, spit them out, and start pooling saliva for 30 seconds. Then, for the next two replicates, the stimulus was not reintroduced, such that the three replicates represented saliva at 30, 60, and 90 seconds after the introduction of stimuli, respectively. All nine falcon tubes for this part of the study were pre-weighed in order to measure the weight of collected saliva.

### 6. Sample disposal and measurements

Pictures of the saliva collection devices and the falcon tubes used in the saliva production rate measurement were taken. The weight and volume of each falcon tube were measured. For the saliva collection devices, volume or the mass of the saliva was measured. Following measurement, all devices and tubes were discarded, and all equipment used was sanitized.

### 7. Statistical analysis of data

For comparing the saliva collection and instruction reading rates for the five devices, one-way ANOVA and Tukey’s HSD Pairwise Comparison were used. For comparing survey responses on instruction, assembly, and collection difficulties among the five devices, one-way ANOVA and Tukey’s HSD Pairwise Comparison were used, by enumerating Likert-scale responses (Very easy, Easy, Fair, Difficult, and Very difficult) to integers 1-5 (respectively, in this order of the responses) where necessary. To compare unstimulated vs. water-stimulated vs. acid-stimulated saliva flow rates pairwise, permutation tests were used. Similarly, within the same condition, to compare female and male salivation rates, permutation tests were done, and the three replicates were averaged. For the relationship between age and saliva flow rate and the relationship between acid-stimulated and water-stimulated saliva flow rates, ordinary least squares regression was used. To compare saliva flow rates at different time points after the introduction of the water and acid stimuli, permutation tests were used in calculating the mean difference and percent change in the mass of saliva collected in 30 seconds. Alpha (maximum value for p-value) was 0.05 (5%) for all tests.

## List of abbreviations

● ICF: Informed Consent Form
● SCA: Saliva Collection Aid
● SVAI: Sample Volume Adequacy Indicator

## Declarations

### Ethics approval and consent to participate

This human-subjects study was deemed exempt by the Caltech IRB.

### Consent for publication

This study contains only deidentified data, and consent was obtained at the time of data collection for each subject.

### Availability of data and materials

The datasets generated and/or analyzed during the current study are available in a GitHub repository, which can be found at: https://github.com/pachterlab/KBP_2023/tree/main. Sections of the dataset containing information about the subjects were removed to protect the privacy of subjects.

### Authors’ contributions

YK and ASB designed the study. YK conducted the experiments and acquired samples. YK and ASB analyzed the samples. YK interpreted the data and was a major contributor in writing the manuscript. All authors read and approved the final manuscript.

## Data Availability

The datasets generated and/or analyzed during the current study are available in a GitHub repository, which can be found at: https://github.com/pachterlab/KBP_2023/tree/main . Sections of the dataset containing information about the subjects were removed to protect the privacy of subjects.

## Acknowledgements

Not applicable

## Additional Files

There is one additional file associated with this manuscript.

● File name: Additional file 1.docx
● Title of data: Supplementary results separating exploratory and confirmatory samples
● Description of data: Additional File 1 contains supplementary results to the main manuscript. The results presented in this file are separated by exploratory (subjects 1 to 30) and confirmatory (31 to 60) samples, whereas the main manuscript contains the results for all subjects.

## Footnotes

1 † We are referring to the funnel and cryovial set-up as SwabSeq, which is the name of a mass-testing platform of COVID-19 testing(8), as this set-up was used in an early implementation of SwabSeq surveillance testing on Caltech Campus. This set-up is not specific to the SwabSeq protocol itself, nor do any of the results from our survey.

## References

1. National Cancer Institute [Internet]. 2011 [cited 2023 Jul 11]. NCI dictionary of Cancer Terms. Available from: https://www.cancer.gov/publications/dictionaries/cancer-terms/def/cushing-syndrome

2. Center for Biologics Evaluation, Research. U.S. Food and Drug Administration. FDA; 2022 [cited 2023 Jul 11]. Information regarding the OraQuick In-Home HIV Test. Available from: https://www.fda.gov/vaccines-blood-biologics/approved-blood-products/information-regarding-oraquick-home-hiv-test

3. Bellagambi FG, Lomonaco T, Salvo P, Vivaldi F, Hangouët M, Ghimenti S, et al. Saliva sampling: Methods and devices. An overview. Trends Analyt Chem [Internet]. 2020 Mar 1;124:115781. Available from: https://www.sciencedirect.com/science/article/pii/S0165993619304182

4. Ott IM, Strine MS, Watkins AE, Boot M, Kalinich CC, Harden CA, et al. Stability of SARS-CoV-2 RNA in Nonsupplemented Saliva. Emerg Infect Dis [Internet]. 2021 Apr;27(4):1146–50. Available from: 10.3201/eid2704.204199

5. Hiremath G, Olive A, Shah S, Davis CM, Shulman RJ, Devaraj S. Comparing methods to collect saliva from children to analyze cytokines related to allergic inflammation. Ann Allergy Asthma Immunol [Internet]. 2015 Jan;114(1):63–4. Available from: 10.1016/j.anai.2014.09.012

6. Kidd S, Midgley P, Lone N, Wallace AM, Nicol M, Smith J, et al. A re-investigation of saliva collection procedures that highlights the risk of potential positive interference in cortisol immunoassay. Steroids [Internet]. 2009 Aug;74(8):666–8. Available from: 10.1016/j.steroids.2009.02.009

7. Tan SH, Allicock O, Armstrong-Hough M, Wyllie AL. Saliva as a gold-standard sample for SARS-CoV-2 detection. Lancet Respir Med [Internet]. 2021 Jun;9(6):562–4. Available from: 10.1016/S2213-2600(21)00178-8

8. Bloom JS, Sathe L, Munugala C, Jones EM, Gasperini M, Lubock NB, et al. Massively scaled-up testing for SARS-CoV-2 RNA via next-generation sequencing of pooled and barcoded nasal and saliva samples. Nat Biomed Eng [Internet]. 2021 Jul;5(7):657–65. Available from: 10.1038/s41551-021-00754-5

9. Affoo RH, Foley N, Garrick R, Siqueira WL, Martin RE. Meta-Analysis of Salivary Flow Rates in Young and Older Adults. J Am Geriatr Soc [Internet]. 2015 Oct;63(10):2142–51. Available from: 10.1111/jgs.13652

10. Froehlich DA, Pangborn RM, Whitaker JR. The effect of oral stimulation on human parotid salivary flow rate and alpha-amylase secretion. Physiol Behav [Internet]. 1987;41(3):209–17. Available from: 10.1016/0031-9384(87)90355-6

11. Gittings S, Turnbull N, Henry B, Roberts CJ, Gershkovich P. Characterisation of human saliva as a platform for oral dissolution medium development. Eur J Pharm Biopharm [Internet]. 2015 Apr;91:16–24. Available from: 10.1016/j.ejpb.2015.01.007

