## Additional File 1 for "Comparative survey-based study of non-invasive saliva collection devices"

### Additional File 1 - Supplementary results

### Device Comparison

<u>Normalized Saliva Collection Rate</u>

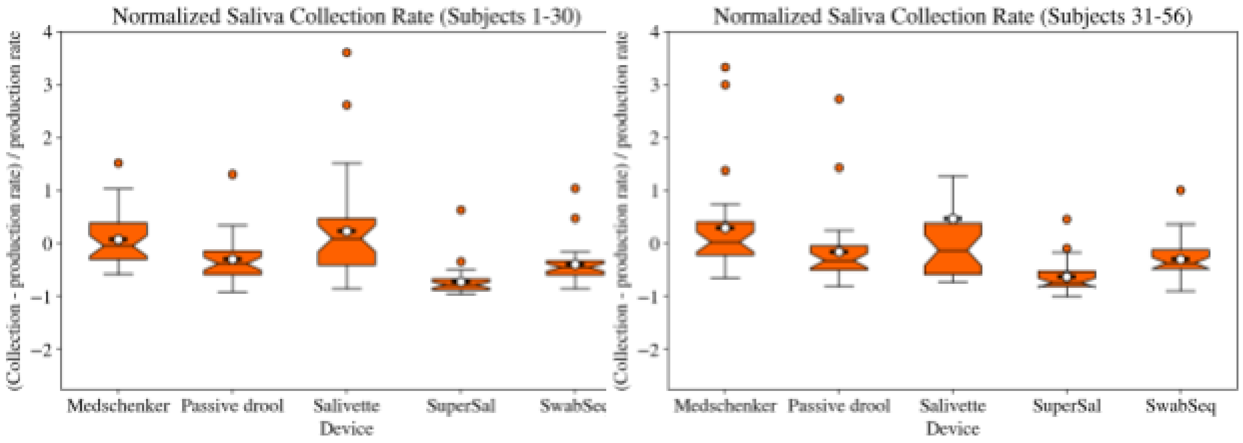


**Figure A1**. Normalized saliva collection rate for each device. Left) Results for exploratory samples, and Right) results for confirmatory samples.

*Exploratory data*

In the exploratory samples with subjects 1 to 30, the average saliva collection rate of each device normalized by each subject's saliva flow rate was ranked Salivette, Medschenker, Passive drool, SwabSeq, and SuperSAL, from highest to lowest. Salivette's and Medschenker's average rates were both above 0, although Medschenker's average rate was close to 0 (0.077578). Rates for Passive drool, SwabSeq, and SuperSAL were all below 0. Out of the five devices, Salivette had the highest average and SuperSAL the lowest. Although Salivette had the highest average, it also had the highest standard deviation and highest interquartile range, meaning that the normalized saliva flow rates for the device were more spread out than other devices. SuperSAL had the lowest average but also had the lowest standard deviation and interquartile range, meaning that the data was quite concentrated in one spot. SuperSAL's average rate was also the farthest away from 0 in magnitude.

*Confirmatory data*

The confirmatory samples with subjects 31-56 showed the same rank in average normalized saliva collection rates as the exploratory samples. Salivette and Medschenker had average rates above 0, and the three other devices had average rates below 0. The average rate for Salivette was observed to be higher than the 75th percentile of the data, possibly due to an outlier with a normalized rate around 13. As with the exploratory samples, Salivette had the highest average and SuperSAL the lowest, Salivette had the highest standard deviation and interquartile range, and SuperSAL had the lowest standard deviation and interquartile range.

**Table A1**. Average normalized saliva collection rate for each device (standard deviation).

| | Exploratory (Subjects 1-30) | Confirmatory (Subjects 31-56) | Combined (Subjects 1-56) |
|---|---|---|---|
| Medschenker | 0.077578 (0.510335) | 0.294602 (0.948411) | 0.178339 (0.747066) |
| Passive drool | -0.302768 (0.443459) | -0.156094 (0.734105) | -0.234669 (0.595062) |
| Salivette | 0.231952 (0.989395) | 0.460246 (2.674389) | 0.337945 (1.94433) |
| SuperSAL | -0.719746 (0.293761) | -0.622109 (0.328029) | -0.674415 (0.311169) |
| SwabSeq | -0.398104 (0.364887) | -0.296488 (0.381923) | -0.350925 (0.372988) |

<u>Normalized Instruction Reading Rate</u>

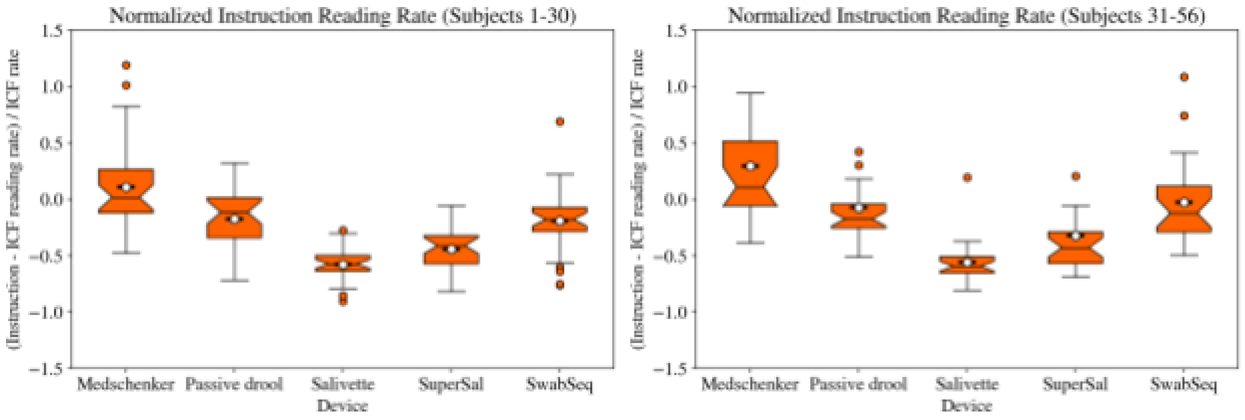


**Figure A2**. Normalized instruction reading rate for each device. Left) Results for exploratory samples, and Right) results for confirmatory samples.

*Exploratory data*

In the exploratory samples with subjects 1 to 30, average normalized instruction reading rates were ranked as follows, from highest to lowest: Medschenker, Passive drool, SwabSeq, SuperSAL, and Salivette. Medschenker, which had the highest normalized rate, had a rate greater than zero on average. However, it had the widest interquartile range and the highest standard deviation. The other devices all had average rates below zero.

*Confirmatory data*

In the confirmatory samples with subjects 31 to 56, the ranks in average normalized instruction reading rates changed to Medschenker, SwabSeq, Passive drool, SuperSAL, and Salivette, from highest to lowest. In contrast to the results from the exploratory samples, SwabSeq's rank was higher than Passive drool's, although the rates for the two devices were very similar in both exploratory and confirmatory samples. In the confirmatory samples, Medschenker had a normalized rate greater than zero, and showed a high variance and wide interquartile range again. Both SwabSeq and Passive drool had normalized rates that were lower than but close to zero, implying that the manuals were almost as easy to read as a plain, text-only document without jargon.

**Table A2**. Average normalized instruction reading rate for each device (standard deviation).

| | Exploratory (Subjects 1-30) | Confirmatory (Subjects 31-56) | Combined (Subjects 1-56) |
|---|---|---|---|
| Medschenker | 0.106586 (0.405584) | 0.294058 (0.612744) | 0.193627 (0.51604) |
| Passive drool | -0.173918 (0.248074) | -0.074995 (0.395087) | -0.127989 (0.32539) |
| Salivette | -0.57668 (0.147904) | -0.559389 (0.188379) | -0.568652 (0.166554) |
| SuperSAL | -0.440979 (0.188856) | -0.319324 (0.528674) | -0.384496 (0.386779) |
| SwabSeq | -0.187781 (0.30257) | -0.02737 (0.370213) | -0.113304 (0.342179) |

Leakage of Saliva

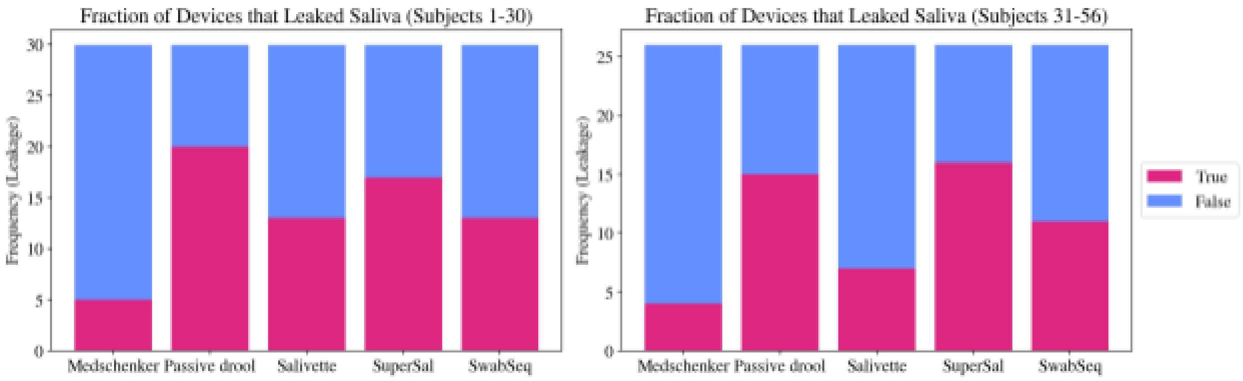


**Figure A3**. Fraction of devices that leaked saliva. Left) Results for exploratory samples, and Right) results for confirmatory samples.

*Exploratory data*

In the exploratory samples with subjects 1 to 30, the fraction of devices with leakage of saliva were ranked as follows, from highest frequency of leakage to lowest: Passive drool, SuperSAL, Salivette and SwabSeq, then Medschenker. Passive drool had the highest fraction of devices that leaked, with more than two-thirds of the devices resulting in leakage. In contrast, Medschenker had the lowest fraction of devices that leaked saliva, with only 5 out of the 30 devices resulting in leakage.

*Confirmatory data*

In the confirmatory samples with subjects 30 to 56, the rankings in fraction of devices with leakage was SuperSAL, Passive drool, SwabSeq, Salivette, and Medschenker. While SuperSAL had the highest occurrence of leakage in the confirmatory samples, with over half of the devices used resulting in leakage of saliva, it was closely followed by Passive drool, which had the highest fraction of leakage in the exploratory samples. Furthermore, while Salivette and SwabSeq were tied in the fraction of devices that leaked in the exploratory samples, SwabSeq had a higher fraction of leakages than Salivette in the confirmatory samples. Although Salivette had around 43% of devices resulting in leakage in the exploratory samples, it had a much lower fraction of 27% in the confirmatory samples. Medschenker continued to show a low fraction of leakages in the confirmatory samples.

**Table A3**. Number of devices that leaked saliva/total number of devices.

| | Exploratory (Subjects 1-30) | Confirmatory (Subjects 31-56) | Combined (Subjects 1-56) |
|---|---|---|---|
| Medschenker | 5/30 | 4/26 | 9/56 |
| Passive drool | 20/30 | 15/26 | 35/56 |
| Salivette | 13/30 | 7/26 | 20/56 |
| SuperSAL | 17/30 | 16/26 | 33/56 |
| SwabSeq | 13/30 | 11/26 | 24/56 |

<u>Survey Results</u>

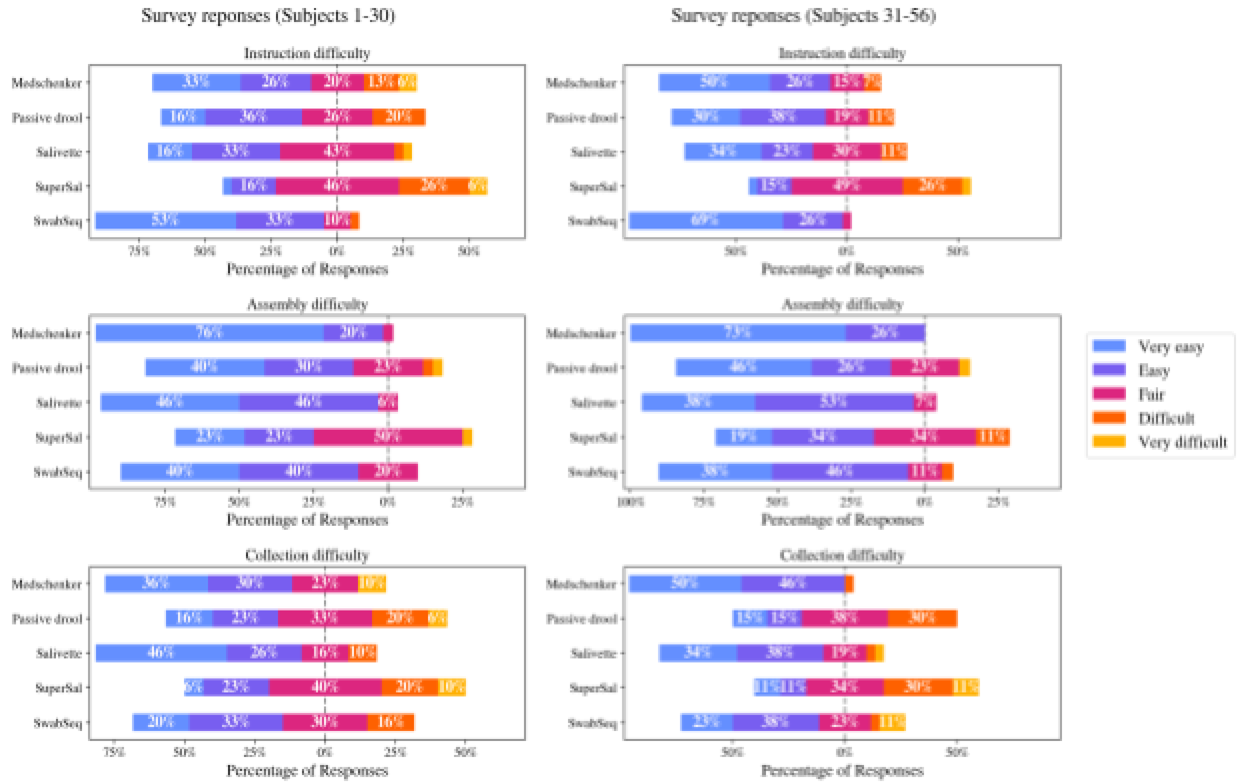

**Figure A4**. Survey results on instruction difficulty (top), assembly difficulty (middle), and saliva collection difficulty (bottom) for each device. Left) Results for exploratory samples, and Right) results for confirmatory samples.

*Exploratory data*

The responses on the difficulty of instructions from the exploratory samples (subjects 1-30) showed that the instructions for SuperSAL were somewhat difficult to read, based on the fractions of “Difficult” and “Very difficult” responses. On the other hand, SwabSeq’s instructions were quite easy to read, with more than 80% of the responses indicating that the instructions were “Easy” or “Very easy” to read, and no responses indicating that the instructions were “Difficult” or “Very difficult”. As for the other three devices–Passive drool, Salivette, and SuperSAL–more responses indicated that the instructions were fairly easy to read.

The responses on the difficulty of device assembly from the exploratory samples (subjects 1-30) for all devices were fairly easy to assemble. Medschenker and Salivette had over 90% of responses indicating “Very easy” and “Easy.” SuperSAL’s assembly difficulty responses had about 50% of responses indicating “Fair” and one response for “Difficult,” and similarly for Passive drool, about 23% of responses indicated “Fair” difficulty and one response each for “Difficult” and “Very difficult.” However, most of the responses for SuperSAL and Passive drool were either “Very easy” or “Easy,” which indicated that the assembly for those devices was mostly easy, except for a few users who found it difficult.

The responses on the difficulty of saliva collection from the exploratory samples (subjects 1-30) showed that Salivette was the easiest to collect saliva with, with about 72% of the responses stating “Very easy” or “Easy” collection. Medschenker also had about 66% of responses indicate “Very easy” or “Easy,” but 10% of the subjects answered that saliva collection was “Very difficult” and no subjects answered “Difficult,” which implies that most users found it easy to collect saliva with Medschenker but there were extreme exceptions. SwabSeq and Passive drool also showed, on average, fairly easy saliva collection. SuperSAL, on the other hand, had an average difficulty of “Difficult,” with a slightly higher portion of responses in “Very difficult” and “Difficult” than responses in “Very easy” or “Easy.”

*Confirmatory data*

The confirmatory samples (subjects 31-56) mostly showed the same trends as the exploratory samples (subjects 1-30), with slight differences in the instruction and collection difficulty ranks. As was observed in the exploratory samples, SwabSeq showed a significantly higher portion of “Very easy” and “Easy” responses than the other devices for instruction difficulty. However, more users in the confirmatory samples found MedSchenker’s and Passive drool’s instructions easier to understand than users in the exploratory samples. In general, subjects in the confirmatory sample had more “Very easy” and “Easy” responses than the subjects in the exploratory sample. The assembly difficulty responses from the confirmatory sample were very similar to the responses from the exploratory sample, with no difference in the rank of the devices in difficulty.

The saliva collection difficulty showed similar trends as the exploratory samples, except that the easiest device to collect saliva with in the confirmatory samples was Medschenker, with 96% of the responses stating “Very easy” or “Easy,” while the easiest device in the exploratory samples was Salivette. Salivette and SwabSeq still had, on average, easy saliva collection. Passive drool had as many responses saying “Difficult” as those saying either “Very easy” or “Easy.” SuperSAL had more responses in “Difficult” and “Very difficult” than in “Very easy” or “Easy,” making it the hardest device to collect saliva with among the confirmatory samples, as was observed in the exploratory samples.

#### Saliva Flow Rates

*Exploratory data*

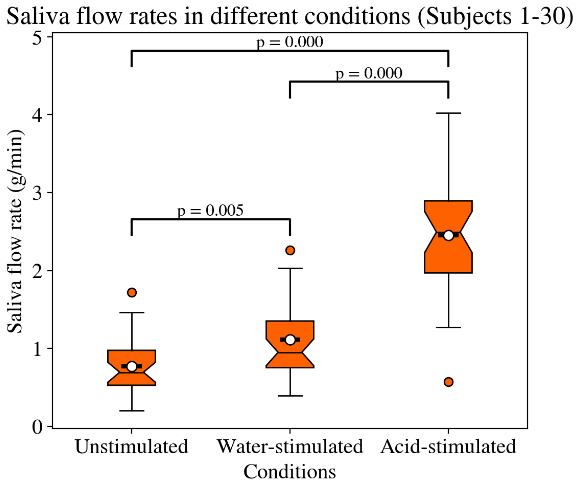


**Figure A5**. Saliva flow rates in unstimulated, water-stimulated, and acid-stimulated conditions for exploratory samples.

The exploratory samples (subjects 1-30) showed statistically significant increases in saliva flow rate in water-stimulated and citric-acid-stimulated conditions compared to unstimulated conditions. The average citric-acid-stimulated saliva flow rate was around 1.68 g/mL higher than unstimulated saliva flow rate and 1.34 g/mL higher than water-stimulated saliva flow rates. The average water-stimulated saliva flow rate was around 0.34 g/mL higher than the average unstimulated flow rate.

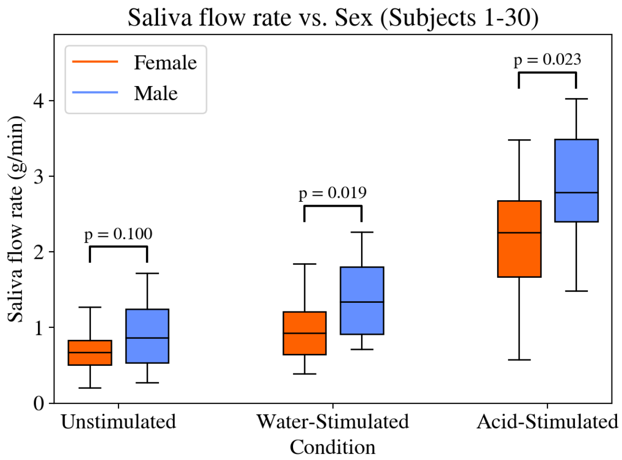

**Figure A6**. Saliva flow rates of exploratory samples in different conditions, separated by sex of subjects.

For unstimulated salivation, there was not a significant difference between male and female saliva flow rates within the exploratory samples. Only in the combined samples were unstimulated saliva flow rates significantly different between females and males. However, for water-stimulated and acid-stimulated saliva flow rates, there were significant differences between male and female saliva flow rates. For water-stimulated saliva, male salivation flow rates were 0.428 g/min higher than those of female subjects. For acid-stimulated saliva, the difference was even greater than that in water-stimulated saliva: male salivation flow rates were 0.690 g/min higher than those of female subjects.

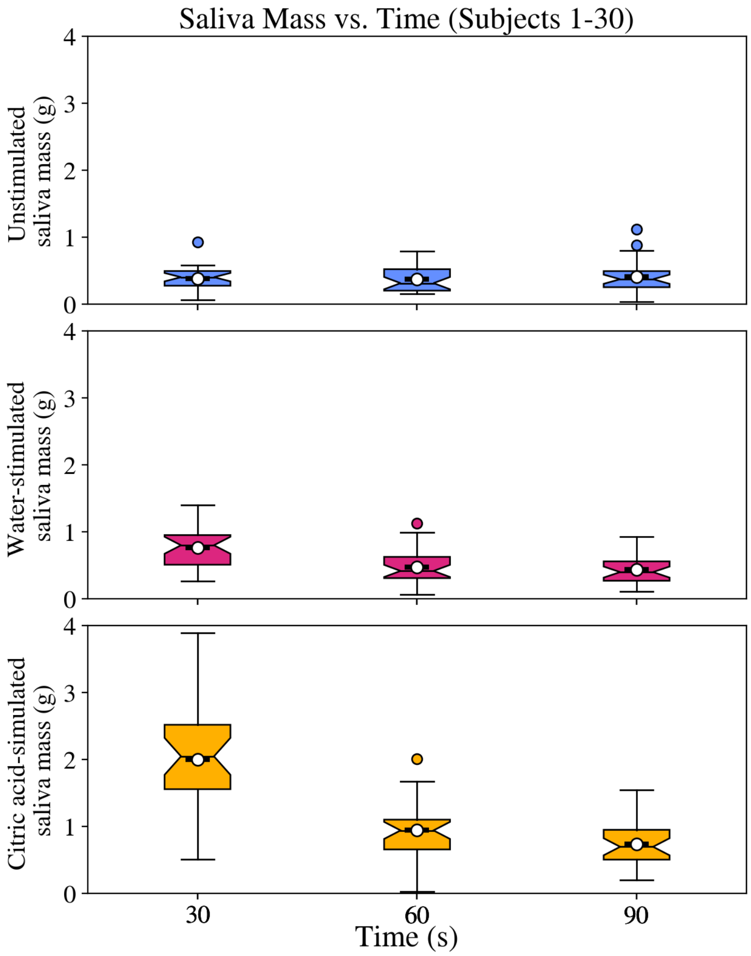

**Figure A7**. Change in saliva mass over time after stimulus introduction for exploratory samples. Stimulus was introduced at t=0, and three samples were collected for each condition, in 30-second intervals

Across the three replicates that were taken at 30 seconds, 60 seconds, and 90 seconds after the introduction of stimuli, respectively, the unstimulated saliva flow rates did not change significantly. Water-stimulated saliva flow rates dropped significantly between 30 seconds and 60 seconds: the difference in mass of collected saliva drops by 0.29 grams between the 30 second and 60 second replicate, which translates to around 38% decrease in mass. However, the difference between the 60 second and 90 second replicates was not significant. For citric-acid-stimulated saliva, significant decreases in saliva flow rates were observed between all three replicates. From the 30 second to 60 second replicate, the collected saliva mass dropped by 1.06 grams, which is equivalent to a 53% decrease. From the 60 second to 90 second replicate, the collected saliva mass dropped by 0.21 grams, which translates to a 22% decrease in mass. These results suggest that water as a stimulus has an effect that lasts up to around 60 seconds after stimulus introduction, whereas citric acid has a longer-lasting effect that is observed even at 90 seconds after stimulus introduction.

*Confirmatory data*

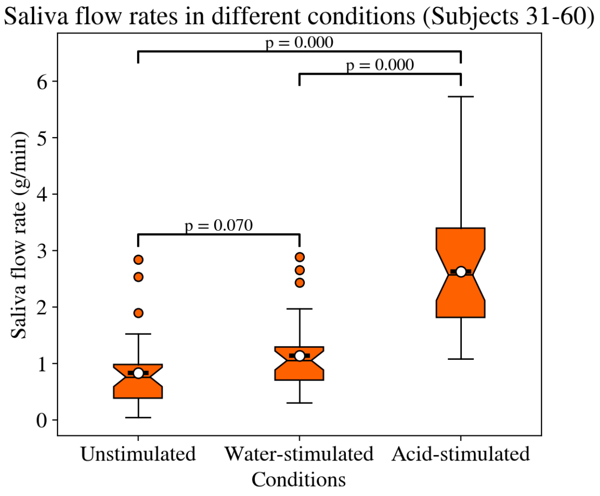

**Figure A8**. Saliva flow rates in unstimulated, water-stimulated, and acid-stimulated conditions for exploratory samples.

The confirmatory samples (subjects 31-60) showed a statistically significant increase in saliva flow rate in citric-acid-stimulated conditions, compared to unstimulated and water-stimulated conditions. The average citric-acid-stimulated saliva flow rate was 1.80 g/mL higher than the average unstimulated saliva flow rate and around 1.49 g/mL higher than the average water-stimulated saliva flow rate. Water-stimulated saliva flow rates were not significantly higher than unstimulated saliva flow rates.

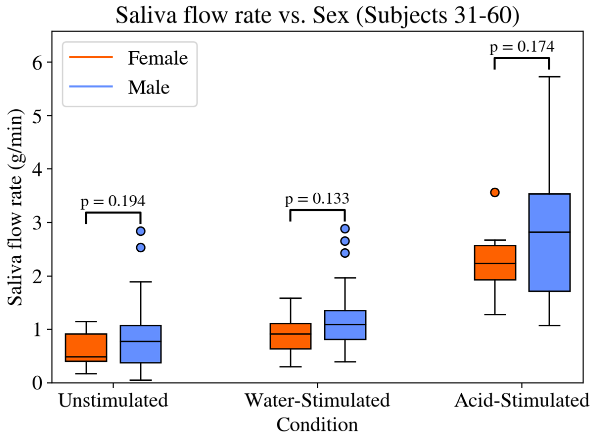


**Figure A9**. Saliva flow rates of exploratory samples in different conditions, separated by sex of subjects.

In contrast to what was observed in the exploratory samples, in the confirmatory samples, there was no significant difference between sexes within the salivation condition. Only in the combined samples were unstimulated saliva flow rates significantly different between females and males.

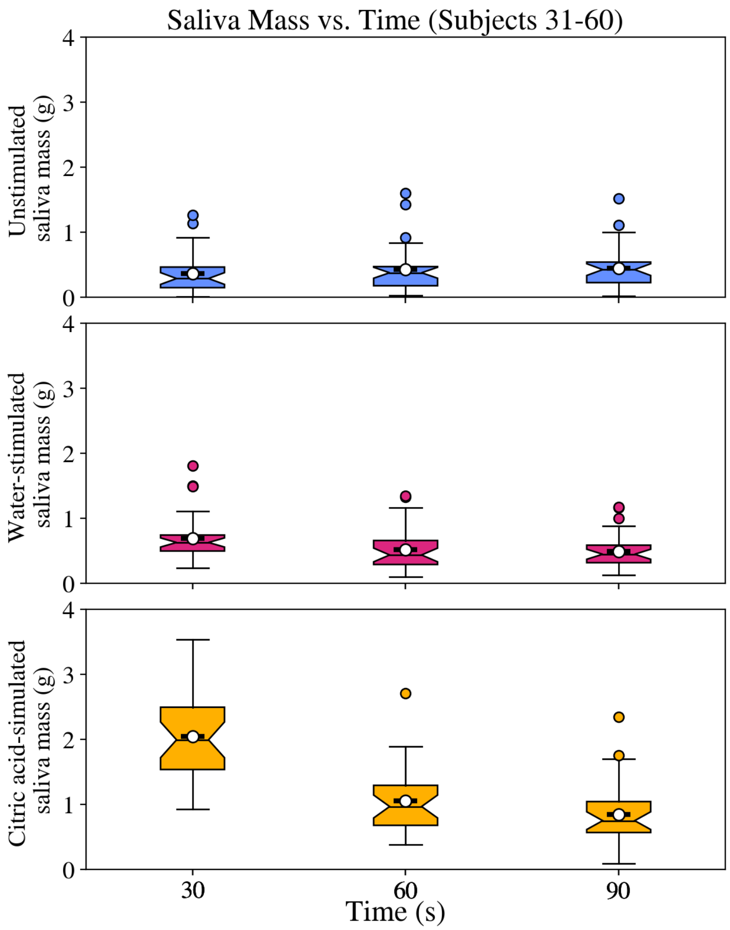


**Figure A10**. Change in saliva mass over time after stimulus introduction for confirmatory samples. Stimulus was introduced at t=0, and three samples were collected for each condition, in 30-second intervals

Within the confirmatory samples (subjects 31-60), unstimulated or water-stimulated saliva flow rates did not change significantly across the three replicates. For citric-acid-stimulated saliva, significant decreases in saliva flow rates were observed only between the 30 second and 60 second replicates. Between these replicates, the collected saliva mass dropped by 0.99 grams, which translates to around 48% decrease in mass. These results, along with the insignificant change in saliva flow rate after water stimulation, contradict the observations made in the exploratory sample which indicated that water indeed increases salivation and has effects that can last up to 60 seconds. The results from confirmatory samples also show that the effects of citric acid may only last for up to 60 seconds, as opposed to the 90 seconds observed in the exploratory samples.

**Table A4**. Average saliva flow rate (g/min) (standard deviation) for each condition and sex

| | | Exploratory (Subjects 1-30) | | Confirmatory (Subjects 31-60) | | Combined (Subjects 1-56) | |
|---|---|---|---|---|---|---|---|
| Condition | Sex | Mean saliva flow rate (g/min) (Standard deviation) | Female - Male mean saliva flow rate (g/min) (p-value) | Mean saliva flow rate (g/min) (Standard deviation) | Female - Male mean saliva flow rate (g/min) (p-value) | Mean saliva flow rate (g/min) (Standard deviation) | Female - Male mean saliva flow rate (g/min) (p-value) |
| Unstimulated | All | 0.772 (0.375) (n=30) | | 0.829 (0.653) (n=30) | | 0.801 (0.529) (n=60) | |
| | Male | 0.911 (0.450) (n=12) | -0.230 (0.0998) | 0.957 (0.789) (n=18) | -0.331 (0.194) | 0.938 (0.665) (n=30) | -0.279 (0.047) |
| | Female | 0.680 (0.295) (n=18) | | 0.626 (0.323) (n=11) | | 0.659 (0.301) (n=29) | |
| Water-stimulated | All | 1.116 (0.492) | | 1.140 (0.636) | | 1.128 (0.564) | |
| | Male | 1.373 (0.540) | -0.428 (0.0188) | 1.282 (0.742) | -0.367 (0.133) | 1.319 (0.660) | -0.385 (0.0066) |
| | Female | 0.945 (0.383) | | 0.915 (0.381) | | 0.934 (0.375) | |
| Citric-acid-stimulated | All | 2.456 (0.839) | | 2.627 (1.043) | | 2.541 (0.942) | |
| | Male | 2.870 (0.845) | -0.690 (0.0228) | 2.809 (1.210) | -0.556 (0.174) | 2.833 (1.063) | -0.626 (0.0078) |
| | Female | 2.180 (0.733) | | 2.253 (0.615) | | 2.207 (0.680) | |
